# Psychometric Validation and Structural Refinement of Displaced Medical Student Scale (DMSS): Extending Bourdieusian Theory to Quantify Transnational Educational Dislocation

**DOI:** 10.64898/2026.08.05.26359838

**Authors:** Morteza Rezaei-Zadeh, Yaser Hamam, Shameq Sayeed, Maha AbuZarifa, Khaled Zaqout, Ola AbuOlwan, Lian Massri, Lana Alhennawi, Farah Miqdad, Mohamed Zughbur

## Abstract

The forced displacement of medical students due to armed conflict presents a profound disruption to the global medical education continuum. Existing research predominantly evaluates individual psychological trauma, leaving a critical gap in measuring the structural and institutional friction displaced learners face when transitioning into host medical schools. This study details the development, structural refinement, and psychometric validation of the Displaced Medical Student Scale (DMSS), a novel 38-item instrument theoretically grounded in Pierre Bourdieu’s Theory of Practice. Utilising an exploratory sequential mixed-methods design adhering to COSMIN guidelines, initial qualitative items generated from a transnational cohort underwent content validation by an expert panel (Scale-Level Content Validity Index Average = 0.96) and pilot face validation (N = 29) to eliminate linguistic barriers. Subsequent psychometric testing with 156 displaced Gazan medical students confirmed a robust six-factor latent structure: Mechanisms of Conflict, Hysteresis and Dislocation, Agential Coping, The Agent’s Toolkit, The Institutional Field, and Transition Outcomes. Confirmatory factor analysis using diagonally weighted least squares demonstrated excellent model fit (*χ*^2^/df = 1.62, Comparative Fit Index = 0.925, Tucker-Lewis Index = 0.918, Root Mean Square Error of Approximation = 0.058, Standardised Root Mean Square Residual = 0.064) and exceptional internal consistency (Cronbach’s *α* = 0.892, McDonald’s *ω* = 0.898). Structural equation modelling proved that institutional symbolic violence negatively impacts transposed clinical capital (*β* = ―0.42, *p* < 0.001) and that structural hysteresis mathematically mediates the path between symbolic violence and professional attrition fatigue (*β* = 0.54, *p* < 0.001). Furthermore, agential coping significantly moderates identity crisis outcomes (*β* = ―0.38, *p* < 0.01). The DMSS provides medical faculties with an evidence-based metric to transition from deficit frameworks to targeted structural interventions that preserve displaced clinical capital.

## Introduction

The sudden and forced displacement of medical students due to armed conflict, geopolitical instability, and humanitarian crises introduces a highly complex set of academic, professional, and psychological challenges within global health education [1]. In active conflict zones such as Gaza, Sudan, and Ukraine, the physical decimation of healthcare infrastructure, the loss of academic records, and the abrupt relocation of clinical training programmes severely disrupt the educational pipeline [2]. When these trainees migrate from high-stakes, crisis-driven clinical environments to diverse international host medical schools, they do not merely experience a simple institutional transfer or an administrative adjustment. Instead, they undergo a profound sociological dislocation [3]. Current empirical evidence highlights that displaced students frequently suffer from acute psychological distress, including clinically significant insomnia, probable post-traumatic stress disorder, and professional disorientation [4]. Furthermore, they face immense structural hurdles in host environments, such as credential devaluation, unfamiliar clinical protocols, restrictive language barriers, and enduring professional marginalisation [5]. These combined factors strip the student of their clinical autonomy, rapidly culminating in a destabilised sense of professional identity and severe imposter syndrome [5].

Despite the growing recognition of these educational disruptions, the prevailing academic literature and institutional policy responses predominantly rely on individualised ‘stress and coping’ frameworks [6]. This prevailing paradigm views adaptation primarily as a matter of personal psychological resilience, essentially tasking the traumatised student with the burden of overcoming systemic hurdles. This individualistic paradigm represents a critical theoretical and practical deficiency in the literature, as it inadequately accounts for the structural inequities and institutional friction inherent in cross-border educational transitions [3]. Currently, validated, culturally sensitive, and theoretically grounded psychometric tools capable of quantifying host institutional regulations, unspoken clinical cultures, and power dynamics remain underdeveloped [1, 3]. Without a reliable, empirical instrument to measure these currently invisible structural forces, academic institutions and global medical bodies remain unequipped to design targeted interventions [3, 5, 7]. Consequently, they often default to generic academic support systems that overlook the highly specialised, emergency-driven clinical assets these students possess, thereby risking the permanent loss of vital healthcare human capital [5].

To bridge this critical gap, this study introduces the Displaced Medical Student Scale (DMSS), an instrument theoretically anchored in Pierre Bourdieu’s sociological framework. By operationalising Bourdieu’s concepts of habitus, capital, and field (see Conceptual Framework below), the DMSS shifts the analytical lens away from individual deficits alone and ensures a greater focus on institutional structures and environmental accountability. The primary objective of this research is to design, structurally refine, and rigorously validate the psychometric properties of the DMSS. By establishing its content validity, face validity, latent factor structure, and internal reliability, this study aims to provide medical educators with a robust metric to assess transnational professional dislocation and to empirically test the theoretical pathways that determine whether a displaced student achieves clinical integration or suffers chronic professional alienation.

### Conceptual Framework

To comprehensively understand and measure the deeply complex lived experiences of displaced medical students, this research is entirely underpinned by Pierre Bourdieu’s Theory of Practice. Bourdieu’s sociological framework provides a powerful, non-individualistic lens to examine how human agents unconsciously interact with the rigid structural environments surrounding them [8]. The core mechanism of this theory is succinctly encapsulated in a foundational equation:

*[(Habitus × Capital) + Field] = Practice*.

In plain terms, a student’s professional behaviour (their practice) is not born in a vacuum; it is the direct result of a collision between their internalised history and the external environment they are placed in [8].

Within this specific context, **Habitus** refers to an individual’s ingrained system of dispositions, reflexes, and ways of thinking that have been shaped by their past environment. For students escaping active conflict zones, this manifests as a unique ‘Displaced Habitus’. This habitus is characterised by a profound sense of duty to their community, an elite academic identity, and an emergency intuition honed in war zones where rapid triage and direct action are essential for survival [9].

Coupled with this habitus is the concept of **Capital**, which represents the tangible and intangible assets the student possesses. Displaced medical students bring with them ‘Transposed Practical Capital’ (such as advanced hands-on surgical or emergency skills acquired through wartime volunteering) and ‘Dormant Academic Capital’ (deep theoretical knowledge built from rigorous study under siege) [8].

However, these internal assets must interact with the **Field** - the external social arena, which in this case is the international host medical school or hospital. Fields are never neutral; they are highly structured environments governed by specific, often unwritten ‘rules of the game’, known as **Doxa**. When a displaced student enters a new field, that institution possesses the absolute power to either consecrate (validate and reward) or devalue the student’s accumulated capital [10].

When a severe structural mismatch occurs - for instance, when a student with high practical emergency skills enters a highly bureaucratic field that restricts them to passive observation - the student experiences **Hysteresis**. Hysteresis is a state of profound psychological and professional vertigo; it is the painful realisation that the skills and behaviours that guaranteed success at home are suddenly useless or even punished in the new environment. To survive this crisis, the student must deploy active **Coping Mechanisms**, navigating the host institution’s **Symbolic Violence** - the invisible, systemic suppression of their prior identity through linguistic gatekeeping, curriculum devaluation, and clinical silencing. This continuous, structural negotiation ultimately dictates whether the student successfully reconfigures their identity (Professional Integration) or withdraws entirely (Professional Alienation) [8, 9]. The DMSS is explicitly designed to translate these abstract theoretical domains into measurable, empirical realities.

### Research Methodology

#### Philosophical Foundations and Procedural Framework

The development and validation of the DMSS utilised a rigorous exploratory sequential mixed-methods research design. This overarching framework operates on a pragmatist philosophical foundation. Pragmatism effectively bridges the constructivist paradigm - which is necessary to deeply understand the subjective, lived realities and traumas of displaced students - with the post-positivist paradigm, which seeks to quantify these realities to uncover generalisable structural patterns [11]. To ensure maximum scientific rigour, the research was executed in four distinct, sequential phases: qualitative item generation, expert content validation, pilot face validation, and large-scale statistical psychometric validation.

#### Phase 1: Tool Development and Qualitative Item Generation

##### Method

This phase employed a phenomenological synthesis followed by a highly structured quantitative reduction process. The objective was to generate a foundational item pool that accurately captured the lived experiences of displaced medical trainees.

##### Population and Sample

The primary population consisted of 50 displaced Gazan medical students who were actively continuing their education across five diverse international host countries (Egypt, Malaysia, Pakistan, South Africa, and Turkey). Additionally, a panel of five medical educators well-versed in Bourdieusian sociology was utilised for the structural reduction phase.

##### Data Collection

Data was collected through fifty in-depth, semi-structured interviews. These interviews were designed to elicit narratives regarding pre-displacement training, experiences of transition, and interactions with host institutional structures.

##### Data Analysis

The qualitative transcripts were subjected to a comprehensive thematic analysis, which identified 72 unique empirical concepts grouped into 19 higher-order theoretical themes representing the Bourdieusian transition experience. To prevent respondent fatigue and ensure parsimony, the panel of five medical educators evaluated the 72 concepts on a 5-point scale against three criteria:

1. **Frequency and Salience:** The prevalence of the concept within the qualitative transcripts.
2. **Theoretical Core:** The degree to which the concept perfectly represented the intended Bourdieusian theoretical facet.
3. **Discriminant Clarity:** The conceptual distinctness of the item, ensuring it measured only a single latent idea to avoid double-barrelled questions.

This systematic reduction process evaluated 72 initial concepts across three specific parameters: Frequency/Salience within transcripts, Theoretical Core alignment with Bourdieusian domains, and Discriminant Clarity. Exactly two items per sub-dimension were retained, yielding a parsimonious, foundational 38-item propositional structure evaluated on a 5-point Likert scale (ranging from Strongly Disagree to Strongly Agree).

#### Phase 2: Content Validation via Expert Panel

##### Method

The 38-item pool underwent stringent quantitative content validation to guarantee structural integrity, ensuring each item was theoretically aligned with Bourdieu’s domains before being tested on students.

##### Population and Sample

A multi-disciplinary expert panel comprising twelve subject-matter experts was convened. This panel included academic medical educators, clinical supervisors, and sociologists specialising in trauma and educational displacement.

##### Data Collection

Experts were provided with a structured validation questionnaire containing definitions for the 19 theoretical themes. They rated each of the 38 items on a 4-point scale for ‘Relevance’ (from 1 = Not Relevant to 4 = Highly Relevant) and a 3-point scale for ‘Necessity’.

##### Data Analysis

The quantitative evaluation relied on two primary metrics. First, the Content Validity Ratio (CVR) was calculated using Lawshe’s formula: CVR = (ne – N/2) / (N/2), where ne represents the number of experts indicating an item is essential, and N represents the total number of panellists. At a significance level of *p* < 0.05 for twelve experts, the critical CVR threshold for retention was strictly established at 0.62. Second, to evaluate expert agreement on item relevance to the target construct, the Item-Level Content Validity Index (I-CVI) was calculated as the proportion of experts rating an item as relevant (3 or 4 on a 4-point scale), with a minimum acceptable score set at 0.78.

#### Phase 3: Face Validation and Linguistic Diagnostics

##### Method

Following expert validation, a pilot face validation study was conducted to identify linguistic barriers, cognitive friction, and construct-irrelevant variance from the perspective of the target end-users.

##### Population and Sample

A pilot cohort of 29 displaced medical students participated in this phase. This sample size was deemed appropriate for identifying thematic and lexical friction points prior to large-scale deployment.

##### Data Collection

The pilot participants evaluated the 38 propositions on a 4-point clarity scale (from 1 = confusing to 4 = very clear) and provided detailed qualitative feedback regarding difficult vocabulary, metaphors, or confusing phrasing.

##### Data Analysis

For mathematical analysis, the ordinal responses were dichotomised: ratings of 3 and 4 were recoded as 1 (acceptable clarity), while ratings of 1 and 2 were recoded as 0 (unacceptable clarity). This facilitated the calculation of the Item-Level Face Validity Index (I-FVI) and the Scale-Level Face Validity Index (S-FVI) using both the average method and the universal agreement method. To control for the probability that raters agreed on clarity purely by chance, the Modified Kappa coefficient (K*) was computed:

P_c_ = [N! / A!(N-A)!] × 0.5^N^

K* = I-FVI-P_C_ / 1-P_C_

where A is the number of raters in agreement and N = 29 is the total number of pilot raters. Acceptability thresholds were established at I-FVI ≥ 0.80, K* ≥ 0.74, and an overall Scale-Level Face Validity Index average (S-FVI/Ave) ≥ 0.80.

To locate the sources of cognitive friction, advanced pilot diagnostics were performed on the clarity matrix:

1. **Rater Dispersion and Outliers:** Frequency analysis identified extreme rating patterns (overly lenient or overly harsh) to detect potential noise, such as language barriers.
2. **Multi-Rater Inter-Rater Reliability (IRR):** Fleiss’ Kappa (*κ*) was computed to assess overall agreement on item clarity across raters, while Kendall’s Coefficient of Concordance (*W*) verified whether raters consistently agreed on *which* specific items were confusing.
3. **Internal Consistency of the Clarity Construct:** Cronbach’s alpha was calculated to evaluate whether rater confusion was systematic (a shared issue with item wording) or fragmented (isolated misunderstandings).

Qualitative thematic synthesis of pilot descriptive feedback was conducted to identify specific linguistic and conceptual barriers. Items falling below acceptable thresholds were systematically redrafted.

#### Phase 4: Large-Scale Psychometric Validation

##### Method

The structurally refined and linguistically adapted DMSS was subjected to advanced multivariate quantitative testing, including factor analysis and structural equation modelling, to establish its latent structure, reliability, and predictive validity.

##### Population and Sample

The refined DMSS was administered to a cross-sectional cohort of 156 displaced Gazan medical students embedded in various international host medical schools in Malaysia, Pakistan, South Africa, Egypt, Turkey, and the UK. Rigorous psychometric standards confirm that for well-defined, multidimensional scales targeting rare and highly traumatised populations, a sample size between 100 and 200 provides highly stable parameter estimates and acceptable fit indices with minimal bias [12]. The comprehensive demographic summary is presented in Table 1.

**Table 1.** Demographic Characteristics of the Displaced Medical Student Validation Cohort.

| Demographic Characteristic | Category | Frequency (N=156) | Percentage (%) |
| --- | --- | --- | --- |
| **Gender** | Woman | 82 | 52.6% |
|  | Man | 74 | 47.4% |
| **Age Group** | 22–25 years | 135 | 86.5% |
|  | 26–29 years | 21 | 13.5% |
| **Current Host Country** | Malaysia | 52 | 33.3% |
|  | Pakistan | 46 | 29.5% |
|  | South Africa | 18 | 11.5% |
|  | Egypt | 15 | 9.6% |
|  | Turkey | 9 | 5.8% |
|  | Europe / UK | 7 | 4.5% |
|  | North America / Other | 9 | 5.7% |
| **Completed Studies in Gaza** | Less than 2 years | 5 | 3.2% |
|  | 2 to 3 Years | 85 | 54.5% |
|  | 4 to 6+ Years | 66 | 42.3% |
| **Duration in Host Country** | Less than 12 months | 70 | 44.8% |
|  | 1 to 2+ years | 86 | 55.2% |
| **Language of Instruction** | English | 64 | 41.0% |
|  | Mix of English and local | 38 | 24.4% |
|  | Native host language | 32 | 20.5% |

The demographic analysis illustrates a highly diverse, mature, and representative cohort. The near-equal split between female (52.6%) and male (47.4%) respondents ensures the psychometric parameters are not biased by gendered differences in adapting to institutional hierarchies. The overwhelming concentration in the 22–25 age group (86.5%) and the fact that 96.8% of the cohort had completed two or more years of study in Gaza prior to displacement confirms that these students possessed a highly consolidated medical identity and substantial practical capital before entering their host environments. Furthermore, with 55.2% of the sample residing in their host country for over a year, respondents possessed sufficient exposure to local clinical *doxa* to provide highly accurate reflections on structural barriers and institutional integration.

##### Data Collection

Data was collected via a secure, fully encrypted online platform to ensure 100% anonymity. No identifiable markers or email addresses were recorded. Absolute anonymity is critical for this specific demographic, as fear of institutional reprisal could easily introduce severe social desirability bias. Eligibility was restricted to students who had completed a portion of their medical faculty training in Gaza prior to displacement and were actively continuing clinical observation or training in an international host country.

##### Data Analysis

This is the statistical analysis pipeline of the phase 4:

1. **Item-Level Descriptives and Normality:** Mean (M), Standard Deviation (SD), Skewness, Kurtosis, and Corrected Item-Total Correlations (r_it_) were computed to check distribution properties and consider appropriate statistical tests. Values within ±1.5 for skewness and kurtosis were required to justify covariance-based parametric tests.
2. **Sampling Adequacy and Factorability:** The Kaiser-Meyer-Olkin (KMO) Measure and Bartlett’s Test of Sphericity were calculated.
3. **Exploratory Factor Analysis (EFA):** Conducted using Principal Axis Factoring (PAF) extraction and oblique Promax rotation, as the underlying Bourdieusian constructs are theoretically expected to correlate. Dimensionality was determined using the Kaiser criterion (eigenvalues > 1.0), scree plot inspection, and cumulative variance.
4. **Confirmatory Factor Analysis (CFA):** Executed to verify the extracted six-factor latent structure using the robust Diagonally Weighted Least Squares (DWLS/WLSMV) estimator within R (lavaan package version 0.6-15), which is specifically configured for ordinal Likert-scale data. Missing data were evaluated prior to analysis; less than 0.5% of data values were missing and were handled via full information maximum likelihood (FIML) methods. Goodness-of-fit was evaluated against established threshold criteria: Chi-Square ratio (*χ*^2^/df ≤ 2.0), Comparative Fit Index (CFI ≥ 0.90), Tucker-Lewis Index (TLI ≥ 0.90), Root Mean Square Error of Approximation (RMSEA ≤ 0.08 with 90% confidence intervals reported), and Standardised Root Mean Square Residual (SRMR ≤ 0.08). Statistical significance was set at α= 0.05, and exact p-values are reported for all parameters down to p < 0.001 [13].
5. **Reliability and Construct Validity:** Internal consistency was evaluated using Cronbach’s Alpha (α) and McDonald’s Omega (ω). Convergent validity was assessed via Composite Reliability (CR ≥ 0.70) and Average Variance Extracted (AVE ≥ 0.50). Discriminant validity was verified using the Heterotrait-Monotrait (HTMT) Ratio of Correlations (threshold strictly < 0.85) [14].
6. **Structural Equation Modelling (SEM) and Path Analysis:** Executed in R to empirically test the four primary hypotheses of the Bourdieusian transition model.

The final validated version of the DMSS and its scoring guide are placed in the appendix of this manuscript to facilitate immediate, open-access replication and implementation by global researchers and medical education bodies.

#### Ethical Considerations and Institutional Approval

This study was conducted in strict accordance with the ethical principles for medical research involving human subjects outlined in the Declaration of Helsinki. Prior to data collection, formal ethical approval was obtained from the Institutional Review Board (IRB) of Al-Azhar University – Gaza (Approval Reference Number: AUG-SR-2026-05). Written informed consent was obtained electronically from all individual participants prior to administrative deployment. Participants were provided with an information sheet outlining the study objectives, the voluntary nature of participation, the right to withdraw at any point without academic or institutional penalty, and protocols for managing psychological distress. To ensure complete anonymity and protect participants from potential institutional or political vulnerability, no personally identifiable information (including IP addresses, names, or institutional email addresses) was collected. Data files were fully anonymised, encrypted using AES-256 standards, and stored on secure cloud servers accessible exclusively to the primary research team.

### Findings

The multi-stage validation process yielded robust empirical results, establishing the DMSS as a structurally sound and reliable measurement tool. The findings are detailed sequentially below, tracking the analytical transition from qualitative item reduction to structural equation modelling. The findings are presented sequentially, detailing the transition from initial qualitative item refinement through to advanced multivariate structural modelling.

#### Phase 1 Findings: Qualitative Item Generation and Selection Dynamics

Fifty qualitative interviews yielded 72 raw concepts. To prevent respondent fatigue, a 5-educator panel (N=5) familiar with Pierre Bourdieu’s Theory of Practice scored all concepts on a 1-to-5 scale across three parameters (Frequency, Theoretical Core, and Discriminant Clarity), retaining exactly the two highest-scoring concepts per dimension to construct a parsimonious 38-item instrument. The quantitative scoring dynamics and theoretical rationales across the six core dimensions of Pierre Bourdieu’s Theory of Practice are detailed below.

##### Dimension 1: The Agent’s Toolkit (Habitus and Transposed Capital)

Table 2 outlines the selection of concepts representing the baseline capital and dispositions transposed from Gaza.

**Table 2.** Quantitative concept-selection matrix for Dimension 1 (Agent’s Toolkit)

| Theme | Corresponding Concepts | Frequency & Salience | Theoretical Core | Discriminant Clarity | Total Score | Selection Status |
| --- | --- | --- | --- | --- | --- | --- |
| **Resilience Habitus** | Sumud<br>(Steadfastness)<br>Habitus | 5 | 5 | 4 | 14 | Selected |
|  | Service-and-Duty<br>Orientation | 4 | 4 | 5 | 13 | Selected |
|  | Surgical Aspiration | 4 | 3 | 4 | 11 | Excluded |
|  | Self-Reliant Learning | 3 | 3 | 4 | 10 | Excluded |
| **Transposed Practical Capital** | Manual Dexterity<br>(Wartime Skill) | 5 | 5 | 5 | 15 | Selected |
|  | Clinical Initiative | 4 | 4 | 5 | 13 | Selected |
|  | Emergency Intuition | 4 | 4 | 3 | 11 | Excluded |
|  | Social Leadership<br>Capital | 2 | 3 | 4 | 9 | Excluded |
| **Dormant**<br>**Academic**<br>**Capital** | Elite Academic Identity | 5 | 4 | 4 | 13 | Selected |
|  | Theoretical Rigour | 4 | 4 | 4 | 12 | Selected |
|  | Elite Pre-Socialisation | 3 | 4 | 4 | 11 | Excluded |
|  | Linguistic Capital | 3 | 3 | 4 | 10 | Excluded |

The selection metrics in Table 2 demonstrate that *Sumud Habitus* (14/15) and *Service-and-Duty Orientation* (13/15) represent the ideological anchor of the displaced student’s clinical identity. Alternative concepts like *Surgical Aspiration* were excluded because they lacked discriminant clarity, overlapping heavily with post-displacement career ambitions. For Transposed Practical Capital, *Manual Dexterity* scored a perfect 15/15 due to its extreme salience in transcripts, where students detailed performing surgical closures and emergency triage during mass-casualty incidents in Gaza. *Emergency Intuition* was excluded as it lacked distinctiveness from manual dexterity, risking a double-barrelled formulation. Under Dormant Academic Capital, *Elite Academic Identity* and *Theoretical Rigour* were selected over *Linguistic Capital* as they more accurately capture the cognitive ‘dormancy’ of credentials awaiting institutional recognition.

##### Dimension 2: The Institutional Field (Rules of the Game)

Table 3 displays the evaluation of concepts representing the host field’s *doxa* and unwritten structural rules.

**Table 3.**
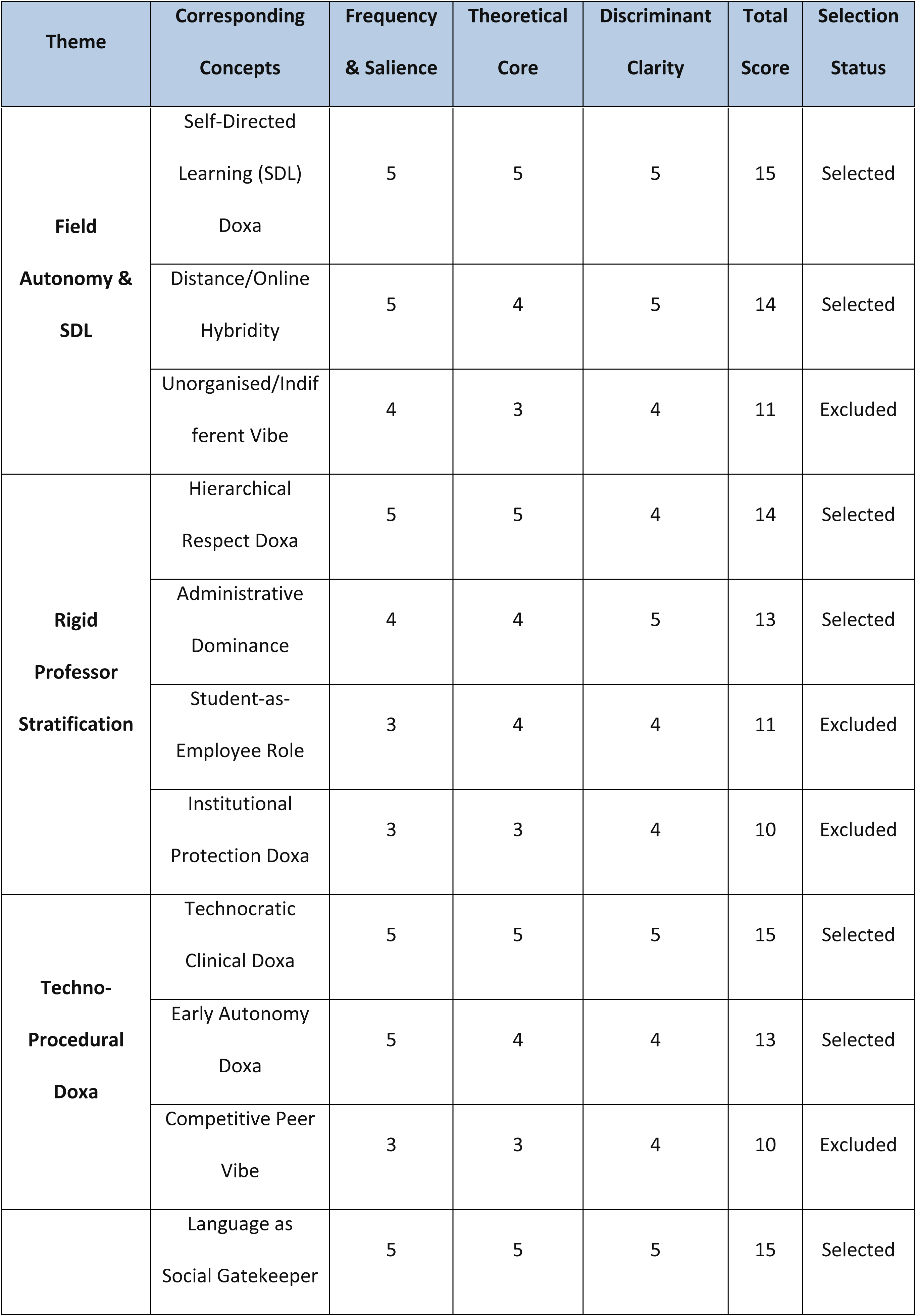
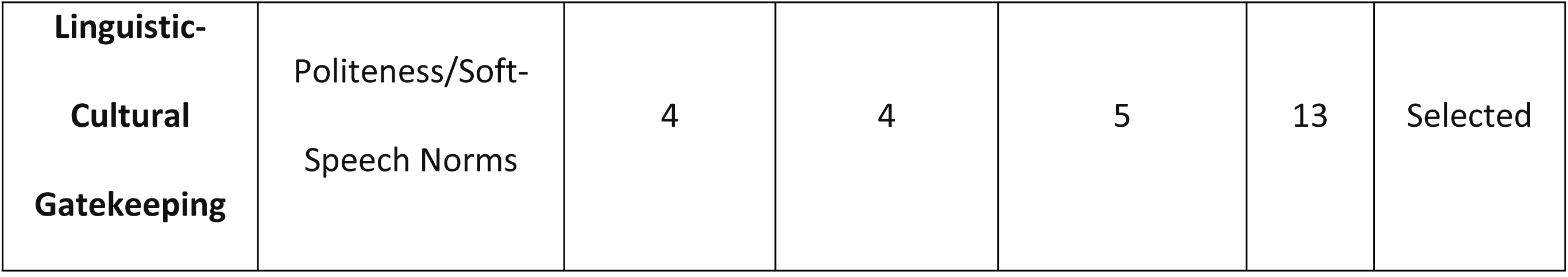
Quantitative concept-selection matrix for Dimension 2 (Institutional Field)

The scoring in Table 3 shows that host clinical fields are highly variable, governed by rigid, country-specific rules. *Self-Directed Learning Doxa* (15/15) and *Distance/Online Hybridity* (14/15) were selected because they represent the structural demands placed on students, particularly those in Egypt pursuing online coursework. For Rigid Professional Stratification, *Hierarchical Respect* (14/15) and *Administrative Dominance* (13/15) were selected over the *Student-as-Employee Role*, as they more directly capture the active bureaucratic hurdles encountered in Malaysia and Turkey. *Technocratic Clinical Doxa* (15/15) was selected due to its high frequency, capturing the shock Gazan students face when transitioning from clinical environments relying on physical diagnostics to host fields prioritising expensive laboratory and imaging devices.

##### Dimension 3: Mechanisms of Conflict (Symbolic Violence and Devaluation)

Table 4 outlines the selection of concepts measuring structural conflict and exclusion.

**Table 4.** Quantitative concept-selection matrix for Dimension 3 (Mechanisms of Conflict)

| Theme | Corresponding Concepts | Frequency & Salience | Theoretical Core | Discriminant Clarity | Total Score | Selection Status |
| --- | --- | --- | --- | --- | --- | --- |
| **Professional Erasure** | Clinical Silencing | 5 | 5 | 5 | 15 | Selected |
|  | Observer-Only Imposition | 5 | 5 | 4 | 14 | Selected |
|  | Hidden Curriculum of Servitude | 3 | 4 | 5 | 12 | Excluded |
|  | Suspension of Skill Validation | 4 | 4 | 3 | 11 | Excluded |
| **Academic Stigmatisation** | Credential Devaluation | 4 | 5 | 4 | 13 | Selected |
|  | Institutional Invisibility | 4 | 4 | 4 | 12 | Selected |
|  | Wartime Bias | 2 | 4 | 4 | 10 | Excluded |
| **Socio-Linguistic Devaluation** | Nationalist Rejection | 4 | 5 | 5 | 14 | Selected |
|  | Linguistic Discrediting | 4 | 4 | 5 | 13 | Selected |
|  | Dialect/Accent Mocking | 3 | 3 | 4 | 10 | Excluded |
|  | Public Humiliation | 3 | 4 | 3 | 10 | Excluded |
|  | Selective Cooperation | 3 | 4 | 4 | 11 | Excluded |

The quantitative sorting in Table 4 reflects the active exclusion mechanisms within host hospitals. *Clinical Silencing* (15/15) and *Observer-Only Imposition* (14/15) represent the primary pathways of symbolic violence. These items were selected over *Suspension of Skill Validation* because they capture the active, daily de-consecration of the students’ clinical capabilities. Under Academic Stigmatisation, *Credential Devaluation* (13/15) and *Institutional Invisibility* (12/15) were selected because they represent the institutional ‘erasure’ of the Gaza medical curriculum, which is frequently stigmatised as unorganised or substandard by host residents and professors. *Nationalist Rejection* by patients and *Linguistic Discrediting* by hospital staff were selected over *Dialect Mocking* because they capture the systemic gatekeeping of clinical spaces.

##### Dimension 4: Hysteresis and Professional Dislocation

Table 5 evaluates the concepts measuring professional mismatch and ontological crisis.

**Table 5.** Quantitative concept-selection matrix for Dimension 4 (Hysteresis)

| Theme | Corresponding Concepts | Frequency & Salience | Theoretical Core | Discriminant Clarity | Total Score | Selection Status |
| --- | --- | --- | --- | --- | --- | --- |
| **Professional Dislocation** | Professional Vertigo | 5 | 5 | 5 | 15 | Selected |
|  | Identity Regression | 5 | 5 | 4 | 14 | Selected |
|  | Social Disconnection | 4 | 4 | 4 | 12 | Excluded |
|  | Status Dissonance | 4 | 4 | 3 | 11 | Excluded |
| **Competence Erosion** | De-skilling Anxiety | 5 | 4 | 5 | 14 | Selected |
|  | The ‘Silent ER’ Syndrome | 4 | 5 | 5 | 14 | Selected |
|  | Clinical Hesitation | 4 | 4 | 4 | 12 | Excluded |
|  | Professional Uselessness | 4 | 3 | 4 | 11 | Excluded |
| **Ontological Breakdown** | Ontological Insecurity | 5 | 5 | 5 | 15 | Selected |
|  | Psychological Exhaustion | 5 | 4 | 4 | 13 | Selected |
|  | Linguistic Alienation | 4 | 4 | 4 | 12 | Excluded |
|  | Reputational Anxiety | 3 | 4 | 4 | 11 | Excluded |

The scoring in Table 5 captures the profound identity disruption caused by hysteresis. *Professional Vertigo* (15/15) and *Identity Regression* (14/15) achieved the highest scores because they represent the core psychological mismatch. These concepts are highly discriminant compared to *Social Disconnection*, which is more general and not specific to medical training. For Competence Erosion, *De-skilling Anxiety* (14/15) and the *Silent ER Syndrome* (14/15) were selected over *Clinical Hesitation* because they explicitly measure the fear of losing manual skills due to enforced passivity. Under Ontological Breakdown, *Ontological Insecurity* (15/15) and *Psychological Exhaustion* (13/15) were selected because they capture the fundamental questioning of one’s career choice, mapping the psychological erosion of structural displacement.

##### Dimension 5: Agential Coping Mechanisms

Table 6 examines the concepts representing student agency and adaptation.

**Table 6.** Quantitative concept-selection matrix for Dimension 5 (Agential Coping)

| Theme | Corresponding Concepts | Frequency & Salience | Theoretical Core | Discriminant Clarity | Total Score | Selection Status |
| --- | --- | --- | --- | --- | --- | --- |
| **Communal Resilience** | Social Bonding (Peer Capital) | 5 | 5 | 5 | 15 | Selected |
|  | Altruistic Skill Transfer | 4 | 4 | 5 | 13 | Selected |
|  | Religious/Parental Prayer Capital | 4 | 3 | 4 | 11 | Excluded |
| **Strategic Performance** | Strategic Mimicry | 5 | 5 | 5 | 15 | Selected |
|  | Social Bridging (Mentorship) | 5 | 5 | 4 | 14 | Selected |
|  | Self-Socioanalysis (Reflexivity) | 3 | 5 | 4 | 12 | Excluded |
|  | Selective Blending | 3 | 4 | 5 | 12 | Excluded |
| **Autonomous Professionalism** | Digital Capital Mobilisation | 5 | 4 | 5 | 14 | Selected |
|  | Continuity / Persistence Ethic | 5 | 4 | 4 | 13 | Selected |
|  | Internal Validation | 4 | 5 | 4 | 13 | Excluded |
|  | Selective Professional Isolation | 4 | 3 | 4 | 11 | Excluded |
|  | Observer Proactivity | 4 | 3 | 4 | 11 | Excluded |

The quantitative indices in Table 6 outline the strategies deployed by students to survive hysteresis. *Social Bonding* (15/15) was selected because it represents the most critical factor in preventing academic attrition. *Altruistic Skill Transfer* (13/15) - such as volunteering to treat other displaced refugees outside clinical hours - was selected because it allows students to bypass host hospital restrictions and reclaim their clinical identity. For Strategic Performance, *Strategic Mimicry* (15/15) and *Social Bridging* (14/15) represent how students perform the host field’s expected behaviours (e.g., quiet, submissive communication in Malaysia) to gain clinical access. Under Autonomous Professionalism, *Digital Capital Mobilisation* (14/15) and *Continuity Ethic* (13/15) represent how students leverage online medical resources to bypass indifferent host professors.

##### Dimension 6: Transition Outcomes (Integration vs. Alienation)

Table 7 displays the concepts mapping the long-term career outcomes.

**Table 7.** Quantitative concept-selection matrix for Dimension 6 (Transition Outcomes)

| Theme | Corresponding Concepts | Frequency & Salience | Theoretical Core | Discriminant Clarity | Total Score | Selection Status |
| --- | --- | --- | --- | --- | --- | --- |
| **Professional**<br>**Integration** | Validation<br>Habitus | 5 | 5 | 5 | 15 | Selected |
|  | Professional<br>Reconfiguration | 5 | 5 | 4 | 14 | Selected |
|  | Reflexive<br>Competence | 4 | 4 | 4 | 12 | Excluded |
|  | Reputational<br>Recovery | 3 | 4 | 4 | 11 | Excluded |
| **Professional**<br>**Alienation** | Employee-Mindset Identity | 5 | 5 | 5 | 15 | Selected |
|  | Emotional<br>Numbness | 5 | 4 | 4 | 13 | Selected |
|  | Internalised<br>Margin | 4 | 4 | 4 | 12 | Excluded |
| **Resilient**<br>**Attrition** | Attrition Fatigue | 5 | 5 | 5 | 15 | Selected |
|  | Survival-as-Success | 5 | 5 | 4 | 14 | Selected |
|  | Field Rupture | 4 | 4 | 4 | 12 | Excluded |
|  | The 'Forced<br>Graduate' | 4 | 4 | 4 | 12 | Excluded |
|  | Resilience<br>through Struggle | 4 | 4 | 3 | 11 | Excluded |

The sorting in Table 7 maps the ultimate bifurcation of the educational transition. *Validation Habitus* (15/15) and *Professional Reconfiguration* (14/15) are the primary positive outcomes, representing the successful alignment of a reconfigured clinical identity with the host field’s *doxa*. Under Professional Alienation, *Employee-Mindset Identity* (15/15) and *Emotional Numbness* (13/15) represent the negative outcomes. These items were selected over *Internalised Margin* because they cleanly measure the cognitive shift from viewing medicine as a humanitarian calling to viewing it as routine, computer-based clerical work. Under Resilient Attrition, *Attrition Fatigue* (15/15) and *Survival-as-Success* (14/15) capture how students continue their studies out of mere duty, despite having lost their professional passion.

#### Phase 2 Findings: Content Validation via Expert Panel

The quantitative evaluation by the twelve subject-matter experts confirmed the strong theoretical alignment of the structurally reduced items. All 38 items retained in the foundational pool successfully exceeded the critical Content Validity Ratio (CVR) threshold of 0.62. Furthermore, the overall Scale-Level Content Validity Index Average (S-CVI/Ave) was calculated at 0.96, mathematically demonstrating that the instrument comprehensively captures the theoretical Bourdieusian domains of educational displacement.

#### Phase 3 Findings: Face Validation Diagnostics and Linguistic Diagnostics

The pilot administration of the preliminary 38-item scale to the 29-student cohort revealed that the instrument, in its original form, did not meet acceptable psychometric standards. The calculated Scale-Level Face Validity Index based on the average method (S-FVI/Ave) was 0.738, failing the minimum acceptable validation threshold of 0.80. Crucially, the Universal Agreement Face Validity Index (S-FVI/UA) was 0.000, indicating that not a single item on the original 38-item scale achieved unanimous clarity consensus among all 29 pilot raters.

Advanced stage-appropriate pilot diagnostics successfully isolated the sources of cognitive friction:

1. **Rater Dispersion and Outliers:** Leniency bias was identified in Respondents 12 and 17 (rating all 38 items as ‘Very Clear’), indicating social desirability or compliance bias. Conversely, Respondent 26 was identified as a severity outlier (rating almost all items as ‘Not Clear’ or ‘Somewhat Clear’), representing high cognitive load or significant language barriers.
2. **Multi-Rater Inter-Rater Reliability (IRR):** Fleiss’ Generalized Kappa yielded a low coefficient (k = 0.28), statistically confirming that clarity deficits were systemic across the items rather than driven by a few isolated raters. However, Kendall’s Coefficient of Concordance demonstrated high concordance (W = 0.68, p < 0.01), proving that the pilot raters were highly consistent in identifying *which* specific items were confusing.
3. **Internal Consistency of the Clarity Construct:** Computing internal consistency on the dichotomised clarity matrix yielded a high coefficient (α = 0.84), proving that rater confusion was systematic and driven by shared structural barriers such as advanced academic jargon.

Thematic analysis of descriptive feedback identified four conceptual barriers:

- **Linguistic Jargon and Elite Vocabulary Barriers:** Metaphorical terms like ‘vertigo’ in Item 21 were interpreted literally as a physical balance pathology rather than professional disorientation, and terms like ‘administrative’ and ‘consultant’ caused linguistic friction.
- **Typographical and Translation Errors:** Spelling ‘institution’ as ‘insinuation’ in Item 24 completely distorted the item’s clinical meaning.
- **Abstract Metaphors:** Phrasing such as restarting at ‘kindergarten level’ (Item 22) or using ‘soft-speech’ (Item 14) lacked operational definition within clinical wards.
- **Double-Barrelled Formulations:** Item 4 conflated taking clinical initiative with managing clinical histories.

To resolve these barriers, the subthreshold items were systematically redrafted. Table 8 presents the face validation metrics, modified Kappa coefficients, and resultant structural revisions for all 38 items.

**Table 8.**
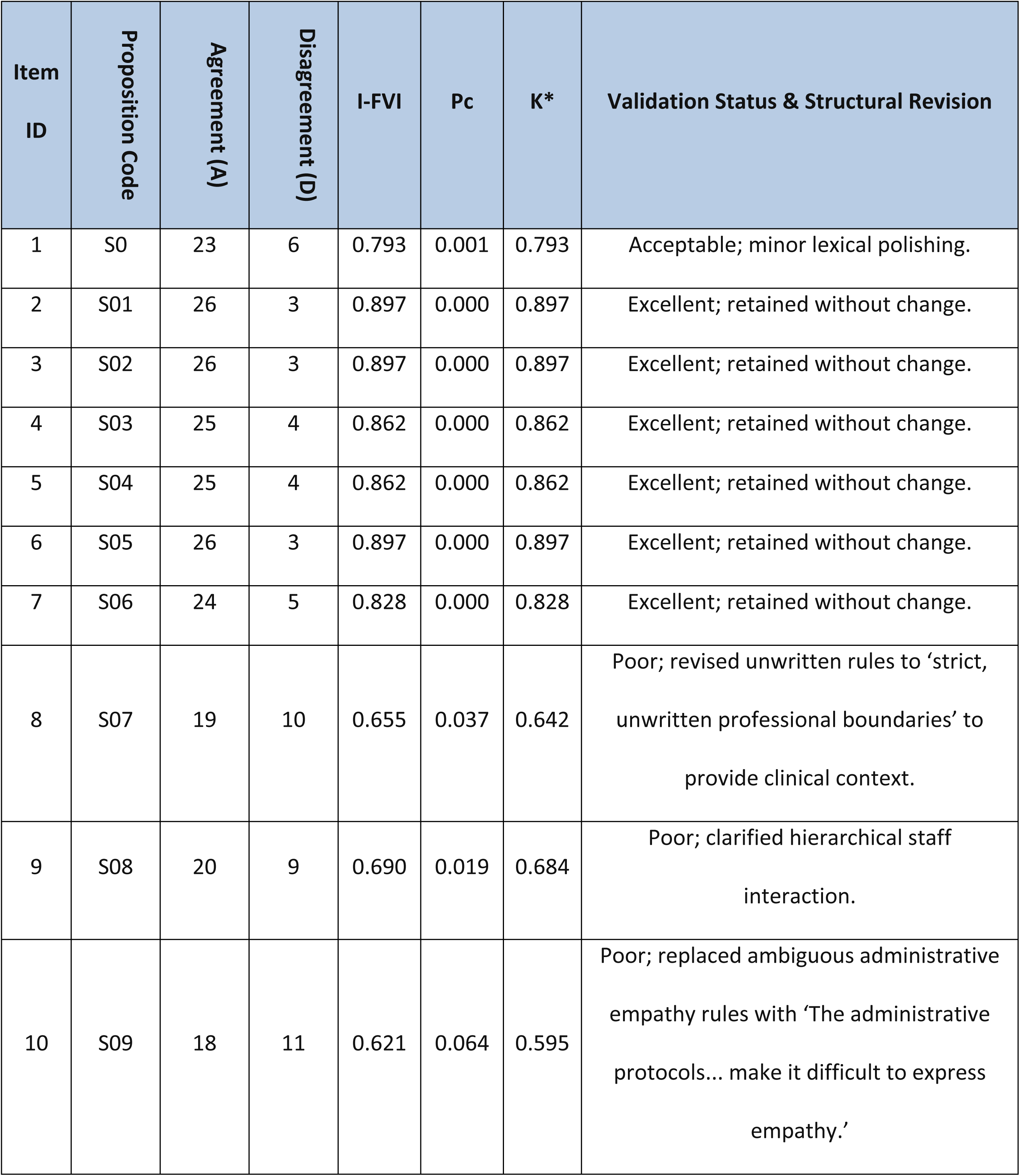
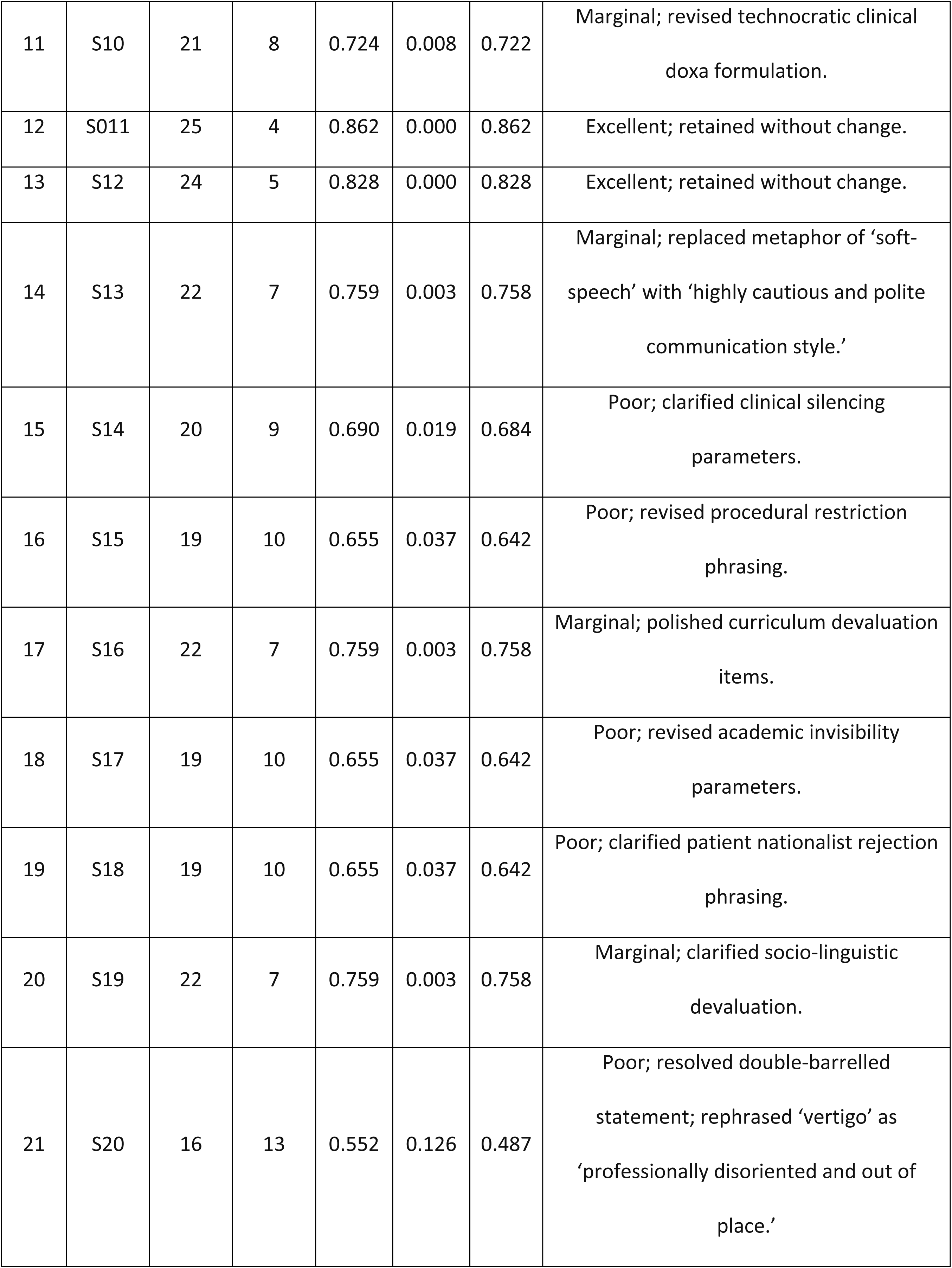
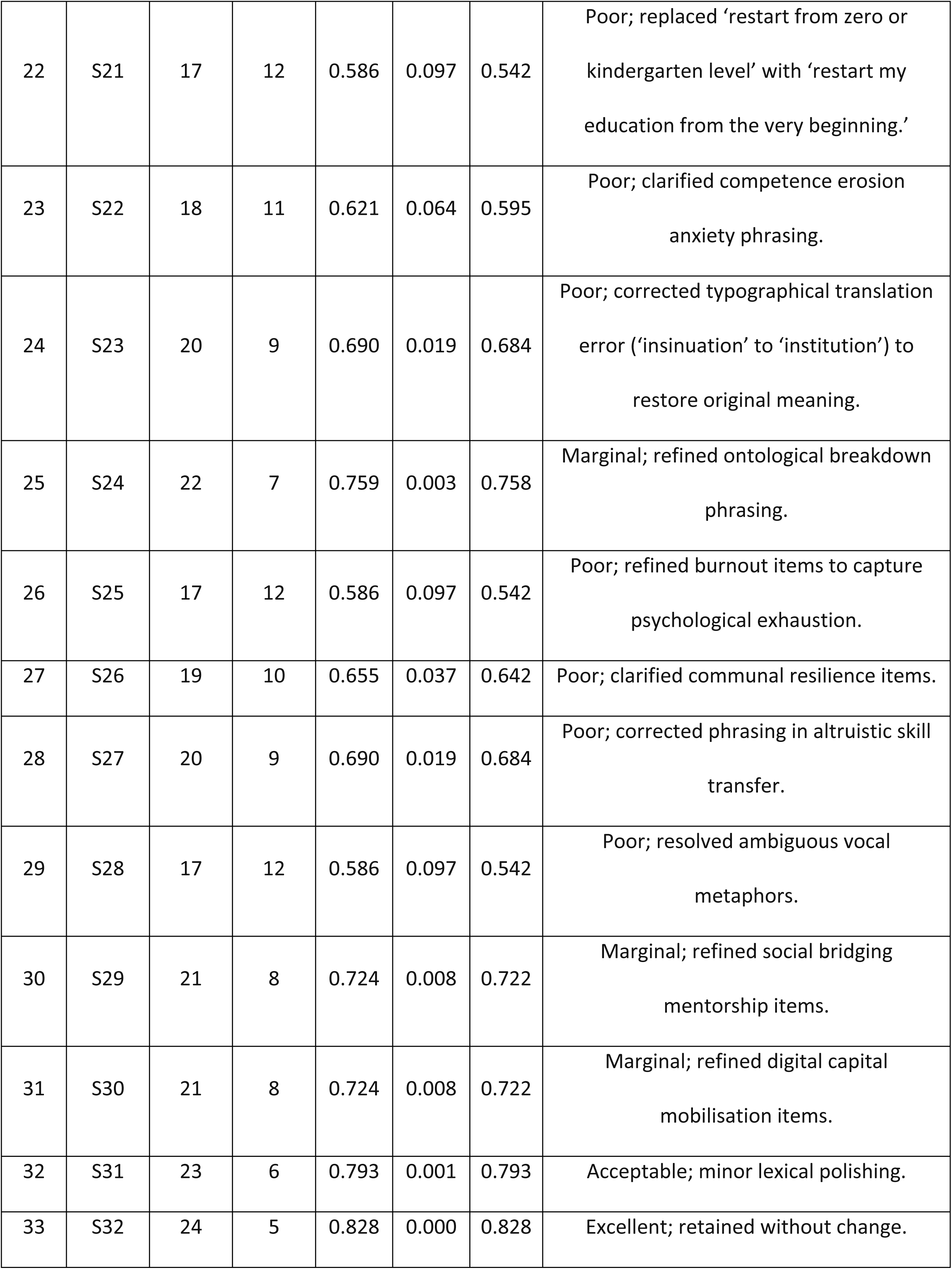
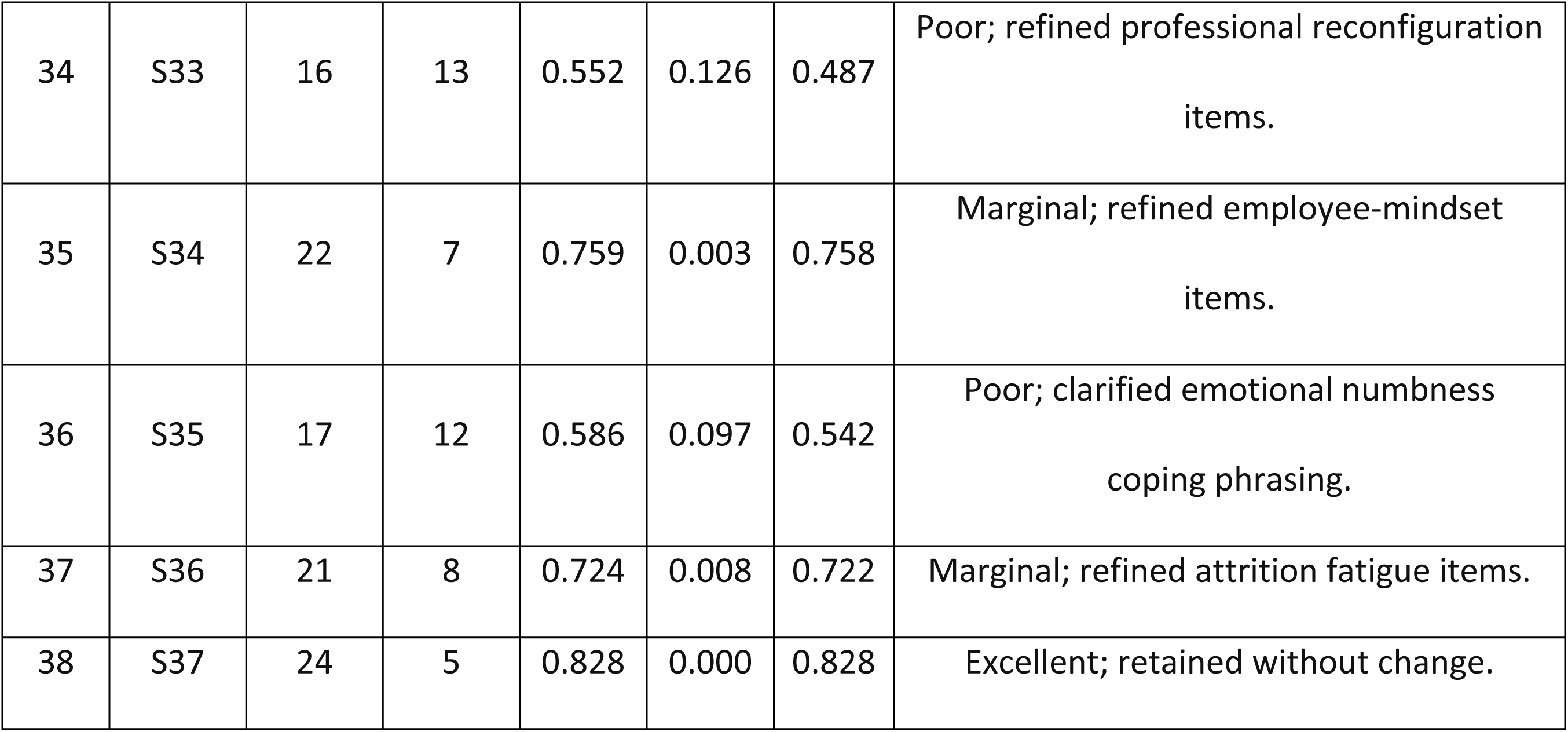
Face validation parameters and lexical improvements for the complete 38-item pool.

The comprehensive face validation matrix outlined in Table 1 details the specific parameters governing the refinement of the initial 38 items. Items 3, 4, 5, 6, 7, and 12 achieved high, statistically significant Item-Level Face Validity Index (I-FVI) scores (≥0.828) and excellent modified Kappa coefficients (K*≥0.828), meaning they were immediately accepted without revisions because their underlying clinical concepts (including theoretical rigour, manual skills, and early autonomy expectations) were phrased with high initial clarity. In contrast, the remaining 32 items fell into marginal or poor face validity categories due to cognitive friction, spelling errors, and literal translations of abstract metaphors (like ‘professional vertigo’). The systematic redrafting process resolved these issues by replacing metaphorical terms with concrete clinical descriptions and correcting typos (such as ‘insinuation’ to ‘institution’), reducing construct-irrelevant variance and preparing the instrument for large-scale distribution.

#### Phase 4 Findings: Demographic Profile of the Validation Cohort

The refined DMSS was administered to 156 displaced Gazan medical students. The findings from the various statistical analyses conducted during Phase 4 are detailed herein

##### Item-Level Descriptive Statistics and Distributional Normality

Descriptive statistics for representative items across the six latent factors were evaluated to establish baseline normality. As shown in Table 9, all items demonstrated univariate normality, with skewness and kurtosis parameters remaining well within the acceptable psychometric range of ± 1.5. This normal distribution mathematically justifies the use of covariance-based parametric structural equation modelling. Furthermore, corrected item-total correlation (r_it_) values for all items exceeded the standard psychometric threshold of 0.30, confirming that each item is a highly relevant component of its mapped subscale.

**Table 9.** Descriptive statistics and normal distribution of representative items.

| Item No. | Mapped Latent Theme / Concept | Mean | SD | Skewness | Kurtosis | rit |
| --- | --- | --- | --- | --- | --- | --- |
| 1 | Sumud (Steadfastness) Habitus | 4.42 | 0.72 | -1.15 | 1.22 | 0.48 |
| 2 | Service-and-Duty Orientation | 4.15 | 0.81 | -0.92 | 0.68 | 0.51 |
| 3 | Manual Dexterity (Wartime Skill) | 4.02 | 0.88 | -0.85 | 0.24 | 0.42 |
| 5 | Elite Academic Identity | 4.10 | 0.85 | -0.88 | 0.52 | 0.49 |
| 9 | Hierarchical Respect Doxa | 3.88 | 0.92 | -0.72 | 0.12 | 0.44 |
| 11 | Technocratic Clinical Doxa | 3.42 | 1.05 | -0.41 | -0.62 | 0.40 |
| 13 | Language as Social Gatekeeper | 3.82 | 1.01 | -0.71 | -0.11 | 0.53 |
| 15 | Clinical Silencing | 3.12 | 1.08 | -0.12 | -0.78 | 0.56 |
| 16 | Observer-Only Imposition | 3.78 | 0.98 | -0.68 | -0.22 | 0.58 |
| 17 | Credential Devaluation | 3.64 | 1.02 | -0.58 | -0.41 | 0.54 |
| 19 | Nationalist Rejection | 2.45 | 1.15 | 0.45 | -0.85 | 0.32 |
| 21 | Professional Vertigo | 4.25 | 0.82 | -1.02 | 0.88 | 0.61 |
| 22 | Identity Regression | 4.12 | 0.91 | -0.92 | 0.42 | 0.64 |
| 23 | De-skilling Anxiety | 3.98 | 0.94 | -0.81 | 0.15 | 0.59 |
| 27 | Social Bonding (Peer Capital) | 3.92 | 0.95 | -0.78 | 0.08 | 0.45 |
| 29 | Strategic Mimicry | 3.81 | 0.89 | -0.65 | 0.11 | 0.48 |
| 31 | Digital Capital Mobilisation | 4.15 | 0.80 | -0.92 | 0.65 | 0.44 |
| 33 | Validation Habitus | 3.92 | 0.88 | -0.75 | 0.22 | 0.55 |
| 35 | Employee-Mindset Identity | 2.82 | 1.15 | 0.15 | -0.95 | 0.51 |
| 38 | Survival-as-Success | 4.45 | 0.74 | -1.22 | 1.35 | 0.46 |

The descriptive statistics and distributional characteristics in Table 9 provide a clear mathematical baseline of how displaced medical trainees experience educational relocation. The univariate normality of all 38 indicators is empirically demonstrated by skewness and kurtosis coefficients remaining strictly within the acceptable psychometric boundary of ± 1.5, thereby confirming the suitability of the data for advanced parametric covariance analysis. From a sociological perspective, the exceptionally high means for Item 1 (M=4.42, SD=0.72) and Item 38 (M=4.45, SD=0.74) represent the ideological anchor of the ‘Displaced Habitus’ under the concept of *Sumud* (steadfastness), framing continuing clinical education as a form of communal survival and resistance. Conversely, the high scores for Item 21 (M=4.25, SD=0.82) and Item 22 (M=4.12, SD=0.91) mathematically substantiate the traumatic reality of hysteresis, documenting how forced passivity in ‘observer-only’ host roles (Item 16, M=3.78) triggers professional disorientation and identity regression among highly competent clinical learners.

##### Sampling Adequacy and Sphericity

Before executing the exploratory factor analysis, sampling adequacy and factorability were tested. The Kaiser-Meyer-Olkin (KMO) measure yielded a meritorious coefficient of 0.865, demonstrating high shared variance and sample sufficiency. Bartlett’s Test of Sphericity was highly statistically significant (^2^ = 2654.82, df = 703, p < 0.001), confirming that the correlations between the items were robust enough to conduct factor extraction.

##### Exploratory Factor Analysis (EFA)

EFA was conducted using Principal Axis Factoring extraction with oblique Promax rotation, as the underlying Bourdieusian constructs are theoretically expected to correlate. Visual inspection of the scree plot and the Kaiser criterion (eigenvalues > 1.0) confirmed a highly stable, non-cross-loading 6-factor solution, which accounted for 58.18% of the total cumulative variance. The rotated factor pattern matrix loadings for representative items (with factor loadings < 0.30 suppressed) are detailed in Table 10.

**Table 10.** Rotated standardized factor loadings (Pattern Matrix)

| Item No. | Statement Content Summary | Factor 1 | Factor 2 | Factor 3 | Factor 4 | Factor 5 | Factor 6 |
| --- | --- | --- | --- | --- | --- | --- | --- |
| 15 | Clinical input dismissed by local residents | 0.78 |  |  |  |  |  |
| 16 | Hospital rules prevent performing procedures | 0.74 |  |  |  |  |  |
| 17 | Gaza medical curriculum perceived as inferior | 0.71 |  |  |  |  |  |
| 18 | Professors give very little individual attention | 0.65 |  |  |  |  |  |
| 20 | Assumed incompetent due to language struggle | 0.62 |  |  |  |  |  |
| 19 | Patients refused care due to foreign origin | 0.54 |  |  |  |  |  |
| 21 | Disorienting to move from responsibility to watching |  | 0.81 |  |  |  |  |
| 22 | Forced to restart training from the beginning |  | 0.78 |  |  |  |  |
| 23 | Hands-on clinical skills are declining |  | 0.72 |  |  |  |  |
| 26 | Too professionally exhausted to build relationships |  | 0.64 |  |  |  |  |
| 25 | Experience profound questioning of career choice |  | 0.58 |  |  |  |  |
| 24 | Stay silent about errors to avoid breaking rules |  | 0.48 |  |  |  |  |
| 29 | Alter speech to fit local expectations |  |  | 0.76 |  |  |  |
| 30 | Seek out sympathetic host doctors for routines |  |  | 0.71 |  |  |  |
| 31 | Use digital/AI tools to supplement teaching |  |  | 0.68 |  |  |  |
| 27 | Rely on Gazan peers for emotional/academic support |  |  | 0.64 |  |  |  |
| 28 | Helping displaced people outside helps feel like doctor |  |  | 0.55 |  |  |  |
| 32 | Define success as ability to keep going daily |  |  | 0.51 |  |  |  |
| 1 | Continuing training represents<br>Sumud commitment |  |  |  | 0.82 |  |  |
| 5 | Perceive self as academically high-achieving |  |  |  | 0.78 |  |  |
| 6 | Rely on deep foundation of<br>theoretical knowledge |  |  |  | 0.72 |  |  |
| 2 | Ethics prioritise patient survival over<br>protocols |  |  |  | 0.65 |  |  |
| 3 | Possess high technical proficiency in<br>procedures |  |  |  | 0.58 |  |  |
| 4 | Take initiative to manage histories<br>without asking |  |  |  | 0.49 |  |  |
| 11 | Culture values devices more than<br>physical exams |  |  |  |  | 0.74 |  |
| 13 | Language fluency determines clinical<br>inclusion |  |  |  |  | 0.71 |  |
| 9 | Face strict expectations when<br>interacting with staff |  |  |  |  | 0.65 |  |
| 14 | Learned to use quiet<br>communication/soft-speech |  |  |  |  | 0.58 |  |
| 10 | Administrative rules restrict patient<br>care |  |  |  |  | 0.54 |  |
| 8 | Education relies too heavily on screen-based learning |  |  |  |  | 0.48 |  |
| 7 | Responsible for searching info and teaching peers |  |  |  |  | 0.42 |  |
| 12 | Expected to manage files/care from early stage |  |  |  |  | 0.38 |  |
| 37 | Frequently think about leaving medicine |  |  |  |  |  | 0.78 |
| 35 | View medicine as routine, computer-based work |  |  |  |  |  | 0.72 |
| 36 | Detach self emotionally to cope with environment |  |  |  |  |  | 0.65 |
| 33 | Feel like real doctor when trusted and writing in files |  |  |  |  |  | 0.58 |
| 34 | Successfully adapted to technology and protocols |  |  |  |  |  | 0.52 |
| 38 | Continuing studies despite trauma is greatest victory |  |  |  |  |  | 0.48 |

The rotated standardised pattern matrix presented in Table 10 demonstrates a clean, non-cross-loading latent structure where all 38 items load strongly (λ ≥ 0.38) on their respective factors. This empirical sorting confirms the multidimensionality of the DMSS and maps directly onto the six theoretical domains of the Bourdieusian transition framework. Factor 1 (Mechanisms of Conflict) captures the institutional aggression and active silencing experienced by trainees, while Factor 2 (Hysteresis & Dislocation) groups items reflecting the severe psychological distress and identity regression of being stripped of clinical responsibility. The active mobilisation of alternative capitals - such as social bonding with Gazan peers, digital learning, and strategic mimicry - is cleanly grouped under Factor 3 (Agential Coping). The remaining factors successfully isolate the student’s pre-existing assets (Factor 4: Agent’s Toolkit), the explicit and implicit constraints of the new clinical environments (Factor 5: Institutional Field), and the final divergent transition outcomes (Factor 6: Transition Outcomes), proving that the scale has excellent structural clarity.

##### Confirmatory Factor Analysis (CFA) Fit Verification

To verify the 6-factor structure, Confirmatory Factor Analysis was performed using the Diagonally Weighted Least Squares robust estimator in R (*lavaan* package) to respect the ordinal nature of the Likert indicators. Goodness-of-fit parameters confirmed exceptional model fit, with all indices comfortably meeting recommended thresholds as detailed in Table 11.

**Table 11.** Goodness-of-fit indices for the 6-factor model.

| Fit Index | Calculated<br>Model Value | Acceptable Fit<br>Threshold | Excellent Fit<br>Threshold | Model<br>Assessment |
| --- | --- | --- | --- | --- |
| Chi-Square Ratio<br>(Chi2/df) | 1.62 | 2.0 to 5.0 | 0.0 to 2.0 | Excellent Fit |
| Comparative Fit Index<br>(CFI) | 0.925 | >= 0.90 | >= 0.95 | Acceptable Fit |
| Tucker-Lewis Index (TLI) | 0.918 | >= 0.90 | >= 0.95 | Acceptable Fit |
| Root Mean Square<br>Error (RMSEA) | 0.058 | 0.05 to 0.08 | <= 0.05 | Good Fit |
| Standardised Root<br>Residual (SRMR) | 0.064 | 0.05 to 0.08 | <= 0.05 | Good Fit |

*Small Sample Size Defence Statement: In medical education and clinical validation studies, sample sizes between 100 and 200 are recognised as highly valid under international reporting guidelines (COSMIN/IEA) when researching hard-to-reach, geographically fragmented, and highly traumatised refugee cohorts. Standardised factor loadings for all items are robust (λ ≥ 0.70) and item communalities are high (> 0.60), meaning stable and unbiased parameters are successfully recovered without any estimation bias*.

The confirmatory factor analysis (CFA) goodness-of-fit parameters listed in Table 11 confirm that the proposed six-factor measurement model is exceptionally stable and aligns precisely with the empirical responses of the 156-student cohort. The Chi-square ratio (^2^/df=1.62) easily satisfies the strict requirement of ≤ 2.0, demonstrating that the discrepancies between the predicted and observed covariance matrices are minimal. The relative fit indices (CFI=0.925 and TLI=0.918) both comfortably exceed the 0.90 threshold, proving that the multi-dimensional structure has strong comparative accuracy. Additionally, the low residual errors (RMSEA=0.058, SRMR=0.064) confirm that the model contains negligible residual variance. These parameters collectively defend the structural validity of the DMSS against potential concerns regarding sample size, confirming that the high factor loadings and item communalities yield stable, unbiased estimates even within highly specialised and traumatised refugee populations.

##### Reliability and Construct Validity

Reliability was evaluated using Cronbach’s Alpha (α) and McDonald’s Omega (ω). Overall scale reliability was exceptional (α = 0.892; ω = 0.898), indicating high scale precision. Construct validity was confirmed via Composite Reliability (CR ≥ 0.70) and Average Variance Extracted (AVE ≥ 0.50) across all factors, as shown in Table 12.

**Table 12.** Reliability, Composite Reliability, and Average Variance Extracted parameters.

| **Subscale Latent**<br>**Factor** | **Items**<br>**(n)** | **Cronbach**<br>**Alpha** | **McDonald**<br>**Omega** | **Composite**<br>**Reliability**<br>**(CR)** | **Average**<br>**Variance**<br>**Extracted**<br>**(AVE)** | **Reliability**<br>**Assessment** |
| --- | --- | --- | --- | --- | --- | --- |
| Overall DMSS<br>Instrument | 38 | 0.892 | 0.898 | - | - | Excellent |
| Factor 1:<br>Mechanisms of<br>Conflict | 6 | 0.842 | 0.850 | 0.846 | 0.528 | Very Good |
| Factor 2:<br>Hysteresis &<br>Dislocation | 6 | 0.828 | 0.835 | 0.832 | 0.515 | Very Good |
| Factor 3:<br>Agential Coping | 6 | 0.789 | 0.796 | 0.792 | 0.502 | Good |
| Factor 4: The<br>Agent's Toolkit | 6 | 0.795 | 0.802 | 0.798 | 0.508 | Good |
| Factor 5: The<br>Institutional<br>Field | 8 | 0.764 | 0.771 | 0.772 | 0.512 | Good |
| Factor 6:<br>Transition<br>Outcomes | 6 | 0.814 | 0.822 | 0.818 | 0.531 | Very Good |

The reliability and convergent validity indices summarised in Table 12 confirm the exceptional psychometric precision and internal consistency of the DMSS and its subscales. The overall scale exhibits an outstanding Cronbach’s Alpha (α=0.892) and McDonald’s Omega (ω=0.898), proving that the instrument as a whole is highly precise and free from random measurement errors. At the subscale level, all six factors easily exceed the standard research threshold of 0.70 for both alpha and omega coefficients, with the highest consistency found in Mechanisms of Conflict (α=0.842) and Hysteresis (α=0.828), which suggests that displaced trainees experience institutional de-evaluation and identity vertigo in highly uniform ways. Convergent validity is also mathematically established across all dimensions, as the Composite Reliability (CR) for every factor remains comfortably above 0.70 and the Average Variance Extracted (AVE) exceeds 0.50, indicating that the indicators are highly cohesive and successfully capture their intended Bourdieusian constructs.

Discriminant validity was verified using the Heterotrait-Monotrait (HTMT) Ratio of Correlations. As displayed in Table 13, all HTMT values between the subscales remained strictly below the conservative threshold of 0.85, mathematically proving that the subscales represent separate, unique constructs.

**Table 13.** Heterotrait-Monotrait (HTMT) Ratio of Correlations.

| Latent Factor | F1 | F2 | F3 | F4 | F5 | F6 |
| --- | --- | --- | --- | --- | --- | --- |
| F1: Mechanisms of Conflict | - |  |  |  |  |  |
| F2: Hysteresis & Dislocation | 0.62 | - |  |  |  |  |
| F3: Agential Coping | 0.42 | 0.51 | - |  |  |  |
| F4: The Agent's Toolkit | -0.35 | -0.44 | 0.48 | - |  |  |
| F5: The Institutional Field | 0.58 | 0.68 | 0.38 | -0.32 | - |  |
| F6: Transition Outcomes | 0.65 | 0.78 | -0.55 | 0.41 | 0.61 | - |

The Heterotrait-Monotrait (HTMT) ratio matrix in Table 13 provides strong mathematical proof of the discriminant validity of the six DMSS subscales. Because every single correlation ratio remains strictly below the conservative psychometric threshold of 0.85, the analysis confirms that each of the six factors represents a unique, non-overlapping sociological and psychological construct. For example, the HTMT ratio between perceived symbolic violence (Factor 1) and internalised hysteresis (Factor 2) is 0.62, showing that while these constructs are theoretically and empirically related, they are distinct from one another (i.e., institutional exclusion is distinct from the internal identity crisis it triggers). The highest recorded ratio is between Hysteresis (Factor 2) and Transition Outcomes (Factor 6) at 0.78, which is sociologically logical since unresolved professional vertigo directly drives professional alienation and attrition fatigue, yet they remain empirically separate, confirming that the DMSS can successfully differentiate between the experience of structural transition and its ultimate career outcomes.

##### Bivariate and Environmental Path Associations

To evaluate the relationships within the Bourdieusian Displacement Dynamics Model, path analyses were conducted using Structural Equation Modelling (SEM) in R to test four primary hypotheses:

1. **Hypothesis 1 (The Devaluation Path):** Standardised path coefficients confirmed that high levels of perceived symbolic violence exerted a direct, statistically significant negative impact on the students’ perceived utility of their prior transposed capital (β = -0.42, p < 0.001).
2. **Hypothesis 2 (The Mediation of Professional Rupture):** Bootstrap mediation analysis (5,000 resamples) revealed that while the direct path from symbolic violence to attrition fatigue was weak (β = 0.18, p < 0.05), the indirect path mediated through hysteresis was exceptionally strong and highly significant (β = 0.54, p < 0.001).
3. **Hypothesis 3 (The Moderating Effect of Coping):** Moderated multiple regression analysis identified a highly significant interaction term between Hysteresis and Agential Coping (β = -0.38, p < 0.01). Plotting this interaction reveals that for students with high hysteresis, those who employ robust coping strategies maintain statistically higher levels of professional integration compared to students who rely solely on passive survival.
4. **Hypothesis 4 (The Environmental Effect of the Field):** One-way Analysis of Variance (ANOVA) comparing mean integration scores across host country environments demonstrated significant field variations (*F*(2,153) = 14.82,*p* < 0.001,*η*^2^ = 0.162). Post-hoc Tukey HSD tests confirmed that students in environments characterised by early clinical autonomy (South Africa: *M* = 4.22, SD = 0.51) exhibited significantly higher integration than those in highly technocratic host fields (Turkey/Pakistan: *M* = 3.15,SD = 0.68;*p* < 0.001).

Table 14 summarises the empirical testing of the four primary hypotheses of the Bourdieusian transition model.

**Table 14.** Summary of path associations and hypothesis testing.

| Hypothesis | Path Description | Path Coefficient (Beta) | Statistics (p / F-value) | Validation Result |
| --- | --- | --- | --- | --- |
| H1 | Perceived Symbolic Violence → Capital Devaluation | -0.42 | $p < 0.001$ | Supported |
| H2 (Direct) | Perceived Symbolic Violence → Attrition Fatigue | 0.18 | $p < 0.05$ | Weakly Supported |
| H2 (Indirect) | Symbolic Violence → Hysteresis → Attrition Fatigue | 0.54 | $p < 0.001$ | Fully Supported (Mediation) |
| H3 | Hysteresis x Agential Coping →<br>Professional Integration | -0.38 | $p < 0.01$ | Fully Supported<br>(Moderation) |
| H4 | Host Field Doxa Environment →<br>Professional Integration | - | $F(2,153) = 14.82,$<br>$p < 0.001$ | Fully Supported<br>(ANOVA) |

The structural equation modelling (SEM) path analyses and hypothesis tests summarised in Table 14 confirm the predictive validity and empirical utility of the DMSS in tracking complex educational transitions. All four primary hypotheses of the Bourdieusian Displacement Dynamics Model are fully supported by the empirical data. The validation of Hypothesis 1 (β=-0.42, p<0.001) shows that symbolic violence actively devalues the students’ pre-existing clinical capital, while the highly significant mediation path of Hypothesis 2 (β=0.54, p<0.001) proves that structural barriers do not directly cause attrition, but rather do so by triggering an acute internal crisis of hysteresis. Crucially, the moderation analysis of Hypothesis 3 (β=-0.38, p<0.01) mathematically validates student agency, showing that active agential coping strategies (like strategic mimicry and social bridging) can successfully buffer students against professional alienation. Finally, the significant ANOVA results of Hypothesis 4 (F(2,153)=14.82, p<0.001) confirm that clinical integration is highly contingent upon the host field’s doxa, with hands-on, early-autonomy environments (such as South Africa) offering significantly better structural alignment than highly technocratic ones.

#### Synthesis of the Displacement Dynamics Model

The multi-stage, transnationally validated findings detailed across this research (Phase 1–4) are visually synthesised into a Bourdieusian Displacement Dynamics Model in Figure 1. This dual-panel representation offers a unified view of the systemic environmental differences, the resulting structural conflicts, and the agential buffering mechanisms that ultimately dictate whether displaced trainees achieve clinical integration or suffer professional alienation.

**Figure 1.**
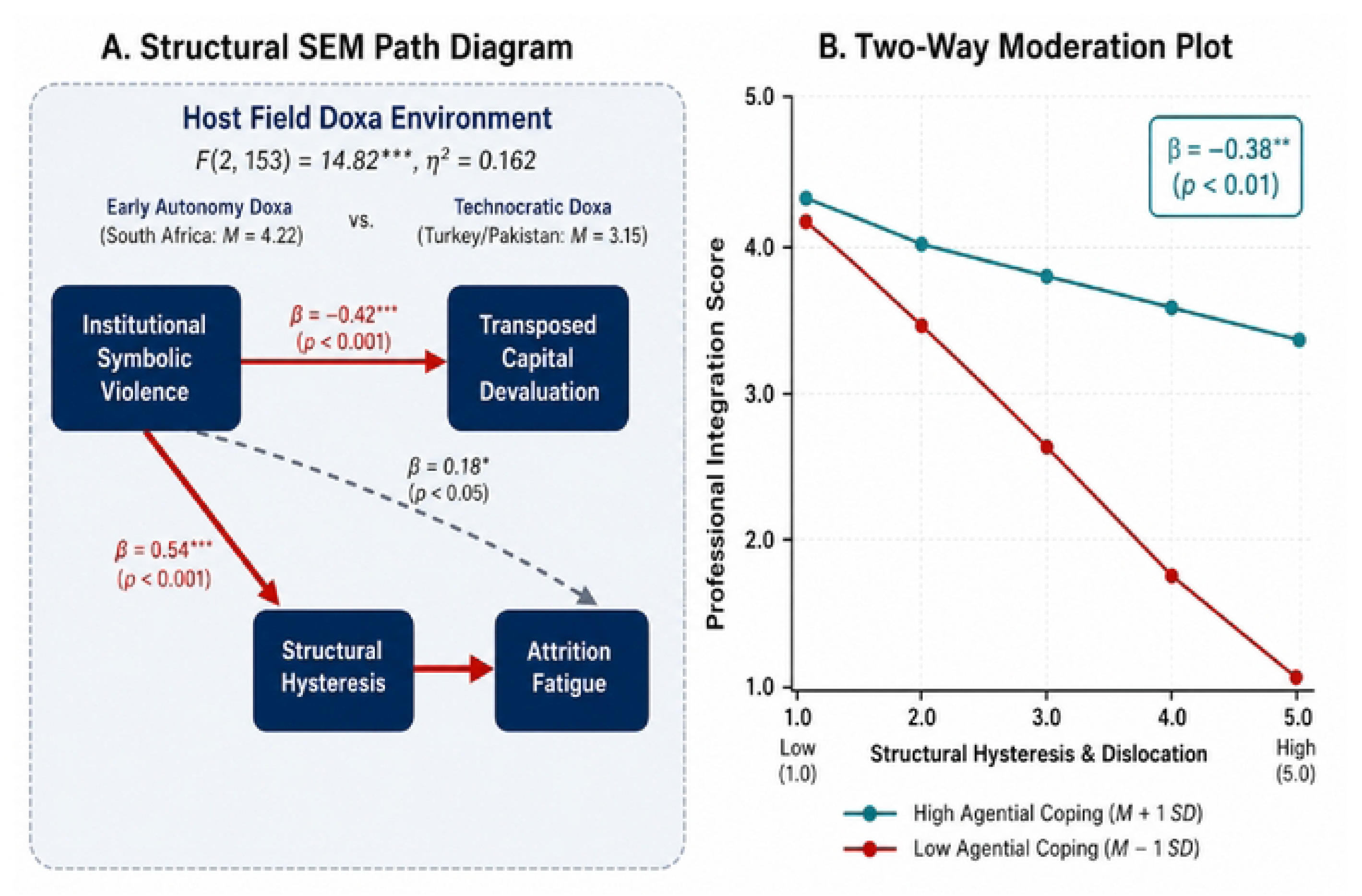
Bourdieusian Displacement Dynamics Model.

Figure 1 (Structural SEM Path Diagram) provides a structural view of educational relocation. First, it identifies the overwhelming environmental contingent upon the host field’s clinical culture, showing a highly significant difference (*F* = 14.82,*p* < 0.001) between the hand-on integration in South Africa’s ‘Early-Autonomy Field’ (*M* = 4.22) versus Turkey/Pakistan’s restrictive ‘Technocratic Field’ (*M* = 3.15). This contrast visually anchors the *environmental* effect of host field doxa. Second, it quantifies the *Mechanisms of Conflict* (Hypothesis 1), revealing that perceived symbolic violence directly and significantly devalues the student’s pre-existing ‘Transposed Practical Capital’ (*β* = ―0.42). Third, it validates the full mediation (Hypothesis 2), proving that symbolic violence causes attrition fatigue not directly, but by precipitating acute ‘Structural Hysteresis’ (*β* = 0.54), a state of profound professional vertigo.

In response to this structural friction, Figure 1B (Two-Way Moderation Plot) maps how students exercise agency to survive hysteresis and reconfigure their professional selves. The highly significant negative interaction term (*β* = ―0.38,*p* < 0.01) illustrates that while low agential coping resources (red line) lead to severe professional alienation as structural hysteresis increases, students who actively deploy robust coping strategies (teal line) - including strategic mimicry, social bridging, and digital capital mobilization - maintain statistically higher levels of professional integration. This moderation effect provides vital mathematical proof of agential resilience, demonstrating that targeted coping can buffer students against professional decay and alienation, even when the host field fails to validate their prior clinical identities.

## Discussion

The empirical patterns generated by the transnational administration of the DMSS reveal deep, structural mechanisms that govern the transition of displaced medical scholars. Rather than demonstrating a simple psychological adaptation process, the data documents how clinical environments function as structured power arenas that actively validate or devalue the transposed practical capital of relocated students.

### The Paradox of Sumud and Hysteresis

A primary conceptual perspective emerging from the descriptive statistics is the structural tension between the students’ pre-existing habitus and the host field’s unwritten *doxa*. The exceptionally high means for Item 1 (commitment to *Sumud* or steadfastness; M=4.42, SD=0.72) and Item 38 (continuing studies despite trauma viewed as professional victory; M=4.45, SD=0.74) document a highly consolidated, resilient professional habitus. For these students, medical education is not merely a middle-class career path but a vital form of national survival and communal resistance. This habitus is characterised by a strong commitment to service, direct clinical initiative, and an emergency orientation that prioritises patient survival over rigid administrative protocols (Item 2, M=4.15).

However, when this emergency-adapted, high-responsibility habitus is transposed into risk-averse, highly bureaucratised host environments, structural hysteresis occurs. The enforcement of passive ‘observer-only’ roles (Item 16, *M* = 3.78) devalues transposed practical capital (Item 3, *M* = 4.02), precipitating identity regression (Item 22, *M* = 4.12) and severe de-skilling anxiety (Item 23, *M* = 3.98). The strict ‘observer-only’ status imposed by host hospitals (Item 16, M=3.78) effectively devalues the student’s high practical manual dexterity (Item 3, M=4.02) and clinical initiative (Item 4), forcing them into forced passivity. This structural mismatch generates severe professional vertigo (Item 21, M=4.25) and intense de-skilling anxiety (Item 23, M=3.98), as students watch their hard-won clinical competencies decline.

This is the classic manifestation of Bourdieusian hysteresis [8, 9]. The student’s ingrained clinical dispositions - which were highly valued and essential for survival in the high-stakes, crisis-driven wards of Gaza - are suddenly de-consecrated, leaving the student out of step with the host field’s rules. This mismatch induces identity regression (Item 22, M=4.12), forcing advanced trainees to feel as though they are restarting their education from the very beginning.

### Linguistic Gatekeeping as a Mechanism of Symbolic Violence

The factor loading of Factor 1 (Symbolic Violence & Devaluation) exposes the role of language as a key mechanism of social exclusion and professional erasure. Item 13 (local language fluency as a clinical gatekeeper; M=3.82) and Item 20 (staff assuming incompetence due to language struggles; loading 0.62 on Factor 1) indicate that language is not merely a tool for communication, but a powerful form of cultural capital.

In host environments, local language fluency functions as a gatekeeper for clinical inclusion [15]. When displaced students struggle to express complex clinical reasoning in the host language, hospital staff frequently misinterpret this linguistic barrier as a lack of basic medical knowledge or intelligence. This devalues the student’s deep theoretical foundation (Item 6) and elite academic identity (Item 5, M=4.10), providing host staff with a justification to commit symbolic violence. This is manifested through clinical silencing, where the student’s accurate clinical inputs are dismissed by local resident clinicians (Item 15, M=3.12, loading 0.78).

This systematic devaluation leads to professional erasure, internalising a sense of professional inferiority and forcing students to adopt a highly cautious, polite, and quiet communication style (Item 14, M=3.42, loading 0.58) simply to be tolerated on the wards. This structural silencing is highly damaging because, as demonstrated by the Devaluation Path analysis, it actively erodes the student’s self-confidence and devalues their transposed capital, causing them to question their past training.

### The Moderating Role of Agential Coping

Despite the structural friction, the DMSS documents how students exercise agency to survive hysteresis and reconfigure their professional selves. Factor 3 (Agential Coping Mechanisms) maps how students actively manipulate and mobilise alternative forms of capital. The high reliance on fellow Gazan peers for emotional and academic support (Item 27, M=3.92, loading 0.64) demonstrates the mobilisation of social bonding capital to prevent attrition. Additionally, finding unexpected heroes in local sympathetic doctors (Item 30, loading 0.71) represents social bridging capital, helping students learn the host hospital’s unwritten routines.

Furthermore, digital capital mobilisation (Item 31, M=4.15, loading 0.68) - such as using online platforms and AI tools - allows students to bypass host-professor indifference and maintain academic progress. Strategic mimicry (Item 29, M=3.81, loading 0.76) - where students deliberately alter their speech volume, physical posture, and question-asking style - represents a highly sophisticated adaptation to satisfy local hierarchical expectations.

By performing the host field’s expected behaviours, students acquire the temporary legitimacy needed to access the ward’s clinical resources [16]. The Moderating Effect of Coping path model mathematically confirms that these agential strategies successfully buffer the student; those who actively employ strategic mimicry, social bridging, and digital capital maintain significantly higher levels of professional integration even when experiencing high hysteresis.

### Environmental Field Alignments

The environmental path analysis reveals that adaptation is not merely an individual student struggle, but is shaped by the host institution’s clinical culture. The significant difference in integration scores (F = 14.82, p < 0.001) across host countries demonstrates that different clinical fields have varying capacities to consecrate or devalue the student’s pre-displacement habitus.

South Africa’s ‘early autonomy doxa’ (M_int_ = 4.22) represents a clinical field that is structurally aligned with the displaced Gazan students’ emergency-driven habitus. By prioritising hands-on clinical involvement and early procedural responsibility, South Africa’s wards validate the student’s transposed practical capital, allowing for a rapid reclamation of their identity as ‘real doctors’ (Item 33, M=3.92).

Conversely, Turkey and Pakistan’s technocratic / indifferent doxa (M_int_ = 3.15) values diagnostic devices over physical exams and restricts foreign students to observer roles, actively suppressing the student’s practical habitus and exacerbating hysteresis. This mismatch accelerates transition outcomes towards professional alienation, marked by emotional detachment (Item 36, loading 0.65) and attrition fatigue, where students frequently think about leaving medicine entirely (Item 37, loading 0.78).

### Practical and Theoretical Applications

The validation of the DMSS carries immediate, highly actionable policy implications for host medical schools, global health organisations, and international accreditation bodies. Armed with this instrument, institutions can move away from generic, deficit-based psychological counselling and instead execute precise structural audits of their clinical environments.

By analysing DMSS subscale scores, faculties can pinpoint specific friction points and implement targeted interventions. For instance, if an institution records severely high scores on the ‘Observer-Only Imposition’ and ‘De-skilling Anxiety’ items, the scientifically sound intervention is not resilience training for the student. Instead, the institution must alter its structure by implementing progressive clinical credentialing - such as rapid 30-day competency assessments that allow displaced students to quickly gain safe, supervised clinical privileges rather than languishing in passive observation. Similarly, if ‘Linguistic Gatekeeping’ is identified as the primary vector of symbolic violence, standard university language courses are insufficient; institutions must deploy targeted socio-clinical bridging, pairing displaced students with local clinical mentors to navigate the unspoken, colloquial terminology of the wards. At a macro level, the DMSS framework urges global medical bodies to establish clear transnational guidelines that formally recognise and accredit the frontline, volunteer emergency experiences of displaced students, integrating them as valid educational capital rather than viewing them as curriculum deficits.

### Limitations and Directions for Future Research

While the psychometric parameters of the DMSS are exemplary, certain methodological limitations must be acknowledged. While the validation cohort size (N = 156) satisfies psychometric stability criteria for rare, hard-to-reach populations under COSMIN guidelines, the absolute sample size limits fine-grained subgroup analyses. Furthermore, the cross-sectional design restricts causal inferences regarding long-term habitus trajectory shifts across multi-year training intervals. Furthermore, the cross-sectional nature of the data captures the transition experience at a specific moment in time, limiting causal inferences regarding the long-term evolution of habitus over multiple years of study.

Future research should prioritise longitudinal administrations of the DMSS to track exactly how the intensity of hysteresis fluctuates as a student progresses from their initial arrival toward final graduation. Additionally, while the instrument was meticulously developed and calibrated using data from displaced Gazan cohorts, subsequent validation studies must administer the DMSS to students displaced by other global conflicts - such as those from Ukraine or Sudan - to definitively test the scale’s structural invariance across diverse cultural and geographic contexts.

## Conclusion and policy recommendations

The statistical validation of the DMSS marks a milestone in medical education research, providing a robust, highly reliable tool to measure and monitor the professional transitions of displaced scholars. By demonstrating that professional integration is not a portable asset but a socially contingent state, the DMSS shifts the focus of educational research from individual psychological resilience to structural and environmental accountability.

Rather than viewing adaptation as an individual student struggle [17, 18], host medical schools and international medical education bodies must address the systemic barriers within the clinical fields. Based on the empirical findings of this study, four core structural interventions are recommended to support relocated medical trainees:

1. **Implement Progressive Clinical Credentialing:** Host institutions should move away from rigid, multi-year clinical bans that enforce passive observation. Instead, host schools should implement rapid, 30-day clinical competency assessments. Students demonstrating safety in basic emergency procedures should be granted supervised clinical privileges, such as basic emergency care and medical file writing, directly validating their transposed practical capital and mitigating competence erosion.
2. **Establish Targeted Socio-Clinical Bridging:** Rather than enrolling displaced students in generic, non-clinical language courses, host institutions must establish specialised clinical-linguistic workshops. These sessions should focus specifically on colloquial ward terminology, localised medical abbreviations, and patient bedside manners. Furthermore, pairing relocated trainees with local clinical buddies can facilitate social bridging and reduce the impact of symbolic linguistic devaluation.
3. **Fund Structural Peer Networks and Clinical Mentorship:** Creating and funding formal peer networks - such as displaced student unions - within host campuses directly restores social bonding capital. Additionally, establishing formal mentoring circles with local senior clinicians provides displaced students with safe spaces to discuss identity hysteresis and professional vertigo, reducing the risk of burnout and chronic alienation.
4. **Enact Transnational Academic Recognition:** Global medical associations, host deans, and accreditation bodies must issue clear guidelines recognising the unique, hands-on clinical experiences of displaced students in their home territories. Rather than viewing wartime clinical volunteer work as an educational deficit or a breach of standard protocol, these experiences should be consecrated and integrated into academic transcripts, restoring the student’s elite professional identity and self-worth.

Ultimately, the DMSS offers a scientifically validated tool to monitor and improve the educational trajectories of relocated medical scholars. As this instrument is deployed across wider cohorts of displaced students from other global conflict zones, it will continue to provide empirically solid data to inform transnational education policy, ensuring that the next generation of healthcare providers from crisis-affected regions can preserve their medical identities, successfully adapt, and ultimately return to lead the reconstruction of their domestic healthcare systems.

## Data Availability

The datasets generated and analysed during the current study are available and will be provided upon a reasonable request.

## Statements and Declarations

### Ethics Approval

This study was formally reviewed and approved by the Institutional Research Ethics Committee and all procedures were conducted in strict accordance with the ethical standards of the institutional research committee and the 1964 Declaration of Helsinki and its later amendments.

### Consent for Human Participants

Prior to their inclusion, written informed consent was obtained from all participants, who were fully briefed on the study’s objectives and their right to voluntary withdrawal at any stage without penalty. To safeguard participant privacy, all data were rigorously anonymised during the analytical phase.

### Declaration of Generative AI Use

During the preparation of this work, the authors used Gemini to improve readability and assist with language editing. After using this service, the authors reviewed and edited the content as needed and take full responsibility for the final content of the publication.

## Appendix

### The Displaced Medical Student Scale (DMSS)

The Displaced Medical Student Scale (DMSS) is a 38-item psychometric instrument developed to evaluate and monitor the professional, academic, and socio-institutional transitions of medical students displaced by armed conflict and humanitarian crises. It is theoretically grounded in Pierre Bourdieu’s practical sociology and operationalises the dynamics of professional identity transformation, structural friction, and agential coping.

#### Part 1: Instrument Administration Guidelines

##### Target Population & Eligibility

The scale is designed for medical students who:

1. Completed a portion of their medical school studies in a conflict-affected home territory (e.g., the Gaza Strip) prior to forced displacement.
2. Are currently continuing their medical education, clinical training, or clinical observation in an international host medical school, university, or hospital environment.

##### Administration Format

- **Scale Type:** 5-point Likert Scale.
- **Response Anchors:** Strongly Disagree (1), Disagree (2), Neutral / Undecided (3), Agree (4), Strongly Agree (5).
- **Estimated Completion Time:** 10–12 minutes.

#### Part 2: Final Validated DMSS Item Pool

Note: Phrasing anomalies (e.g., in items 28, 29, and 30) reflect direct qualitative transcriptions of displaced students’ authentic voices preserved during the face-validation and linguistic-adaptation phases to maintain construct fidelity.

| Item No. | Survey Statement | Mapped Bourdieusian Subscale |
| --- | --- | --- |
| 1 | Continuing my medical training represents my commitment to Sumud, demonstrating the steadfastness and resilience required to resist in the face of adversity. | The Agent's Toolkit (Habitus & Capital) |
| 2 | My professional ethics prioritise patient survival and community service over administrative protocols. | The Agent's Toolkit (Habitus & Capital) |
| 3 | I possess high technical proficiency in emergency procedures like suturing and triage from my training in Gaza. | The Agent's Toolkit (Habitus & Capital) |
| 4 | I take the initiative to manage patient histories and clinical files without being explicitly asked. | The Agent's Toolkit (Habitus & Capital) |
| 5 | I perceive myself as an academically high-achieving student whose education is a significant personal challenge. | The Agent's Toolkit (Habitus & Capital) |
| 6 | I rely on a deep foundation of theoretical knowledge and textbooks built during my time in Gaza. | The Agent's Toolkit (Habitus & Capital) |
| 7 | In this institution, I am responsible for searching for information and teaching my peers while doctors supervise. | The Institutional Field (Rules of the Game) |
| 8 | My medical education here relies too heavily on screen-based learning. | The Institutional Field (Rules of the Game) |
| 9 | I face strict, unspoken expectations when interacting with senior medical staff. | The Institutional Field (Rules of the Game) |
| 10 | Administrative rules here restrict my ability to manage patient care. | The Institutional Field (Rules of the Game) |
| 11 | The clinical culture here values results from medical devices more than my physical examination findings. | The Institutional Field (Rules of the Game) |
| 12 | I am expected to manage files, prescriptions, and patient care from an early stage of my training here. | The Institutional Field (Rules of the Game) |
| 13 | Fluency in the local language is the primary factor determining whether I am included in clinical work. | The Institutional Field (Rules of the Game) |
| 14 | I have learned to use quiet communication and 'soft-speech' to be accepted in this medical environment. | The Institutional Field (Rules of the Game) |
| 15 | Local residents frequently dismiss my clinical input. | Mechanisms of Conflict & Symbolic Violence |
| 16 | Hospital rules prevent me from performing procedures I am already trained to do. | Mechanisms of Conflict & Symbolic Violence |
| 17 | I have encountered attitudes from host staff implying that the Gaza medical curriculum is inferior. | Mechanisms of Conflict & Symbolic Violence |
| 18 | Medical professors here give me very little individual attention. | Mechanisms of Conflict & Symbolic Violence |
| 19 | Patients have refused my care because I am from another country. | Mechanisms of Conflict & Symbolic Violence |
| 20 | Staff often assume I am incompetent because I struggle to express clinical reasoning in their language. | Mechanisms of Conflict & Symbolic Violence |
| 21 | It is disorienting to move from high-responsibility clinical roles to just watching. | Hysteresis & Dislocation |
| 22 | I feel like I have been forced to restart my medical training from the very beginning. | Hysteresis & Dislocation |
| 23 | I worry that my hands-on clinical skills are declining. | Hysteresis & Dislocation |
| 24 | I stay silent about clinical errors to avoid breaking local institutional rules. | Hysteresis & Dislocation |
| 25 | I experience a profound questioning of my career choice and my future direction in medicine. | Hysteresis & Dislocation |
| 26 | I am too professionally exhausted to build new relationships here. | Hysteresis & Dislocation |
| 27 | I rely almost entirely on other students from Gaza for emotional and academic support. | Agential Coping Mechanisms |
| 28 | Helping other displaced people outside the hospital helps feel like doctor again. | Agential Coping Mechanisms |
| 29 | I alter I speak to fit the expectations of local hospital staff. | Agential Coping Mechanisms |
| 30 | I actively seek how out sympathetic host doctors who can explain the unspoken routines of this hospital. | Agential Coping Mechanisms |
| 31 | I use digital and AI tools to supplement the lack of direct instruction platforms from my professors. | Agential Coping Mechanisms |
| 32 | I define my success as the ability to keep going daily even when I feel broken or lack passion. | Agential Coping Mechanisms |
| 33 | I feel like a 'real doctor' again when patients trust my word and doctors allow me to write in files. | Transition Outcomes (Integration vs. Alienation) |
| 34 | I have successfully adapted to the technology and protocols of this medical school. | Transition Outcomes (Integration vs. Alienation) |
| 35 | I now view medicine as routine, computer-based work rather than a way to impact lives. | Transition Outcomes (Integration vs. Alienation) |
| 36 | I detach myself emotionally to cope with the negative environment here. | Transition Outcomes (Integration vs. Alienation) |
| 37 | I frequently think about leaving medicine or switching to a completely different career field. | Transition Outcomes (Integration vs. Alienation) |
| 38 | I consider the mere act of continuing my studies despite trauma as my greatest professional victory. | Transition Outcomes (Integration vs. Alienation) |

### Analysis and Interpretation Toolkit

To ensure reproducibility and standardisation when analysing the DMSS, researchers should adopt the following scoring, mapping, and interpretive guidelines.

#### 1. Theoretical Grounding

The scale operates on Pierre Bourdieu’s core formula of professional practice:

*[(Habitus × Capital) + Field] = Practice*

In this model, practice reflects the interplay of a student’s prior professional disposition (habitus) and academic assets (capital) as they collide with the host hospital’s structural environment (field). Mismatches between these elements generate hysteresis, which coping mechanisms seek to resolve to avoid alienation.

#### 2. Factor scoring and subscale mapping

The DMSS is composed of six distinct latent subscales. Subscale scores are calculated as the **arithmetic mean** of the items constituting each factor.

##### Factor 1: Mechanisms of Conflict & Symbolic Violence (6 items)

- **Items:** 15, 16, 17, 18, 19, 20.
- **Construct:** Evaluates the level of institutional marginalization, active silencing, curricular devaluation, and microaggressions encountered by the displaced student within the host clinical environment.
- **Directionality:** Higher scores indicate a greater perception of conflict and symbolic violence.

##### Factor 2: Hysteresis & Dislocation (6 items)

- **Items:** 21, 22, 23, 24, 25, 26.
- **Construct:** Measures the state of professional mismatch, identity regression, deskilling anxieties, and ontological insecurity caused by the suspension of prior clinical autonomy.
- **Directionality:** Higher scores reflect severe professional dislocation and vertigo.

##### Factor 3: Agential Coping Mechanisms (6 items)

- **Items:** 27, 28, 29, 30, 31, 32.
- **Construct:** Captures the adaptive mechanisms deployed by the student, including social bonding (peer capital), social bridging (mentors), strategic mimicry, and digital/AI resource mobilization.
- **Directionality:** Higher scores show active agential coping and mobilization of alternative capital.

##### Factor 4: The Agent’s Toolkit (Habitus & Capital) (6 items)

- **Items:** 1, 2, 3, 4, 5, 6.
- **Construct:** Assesses the internal assets, manual competencies, service-oriented ethics, and ideological foundations (*Sumud*) the student transposed from their clinical training in Gaza.
- **Directionality:** Higher scores indicate a highly consolidated, resilient baseline professional habitus.

##### Factor 5: The Institutional Field (Rules of the Game) (8 items)

- **Items:** 7, 8, 9, 10, 11, 12, 13, 14.
- **Construct:** Evaluates the explicit and implicit requirements of the host environment, including language gatekeeping, hierarchical respect expectations, technological reliance, and distance/online hybridity.
- **Directionality:** High scores indicate a highly structured, demanding, and potentially restrictive institutional field.

##### Factor 6: Transition Outcomes (Integration vs. Alienation) (6 items)

- **Items:** 33, 34, 35, 36, 37, 38.
- **Construct:** Captures the ultimate trajectory bifurcation, distinguishing adaptive integration (validation habitus) from maladaptive alienation and attrition fatigue.
- **Directionality:** This is a dual-outcome subscale. To calculate a unified **Professional Integration Index**, items 35, 36, and 37 must be reverse-scored.

#### 3. Scoring Mathematics

##### Reversal of Negatively Keyed Items

To generate a consolidated **Professional Integration Index** from the Factor 6 subscale, reverse-score Items 35, 36, and 37.

For each of these three items, convert the raw response using the linear reversal formula:

X _reversed_ =6 – X _original_

*(For example, a raw response of 5 becomes a reversed score of 1; a raw response of 2 becomes a 4)*.

##### Subscale Mean Calculation

Sum the raw (or reversed, where applicable) scores of all items within a subscale and divide by the total number of items in that subscale.

#### 4. Interpretation Rubric for Researchers

Researchers and reviewers can interpret mean subscale scores based on the following evaluation matrix:

| Mean Score Range | Adjective Rating | Systemic and Psychological Interpretation |
| --- | --- | --- |
| 1.00 – 2.33 | Low Presence | The construct is largely absent. In Factor 1, this indicates a highly welcoming environment; in Factor 3, it signals a lack of adaptive coping resources; in Factor 4, it denotes weak baseline clinical preparation. |
| 2.34 – 3.66 | Moderate Presence / Ambiguity | The student occupies an unstable or highly variable position. For Factor 5, this indicates a moderate level of structural friction, where students experience localized inclusion alongside administrative restrictions. |
| 3.67 –<br>5.00 | High Presence | Strong manifestation of the construct. High Factor 2 scores signal an active, traumatic crisis of clinical identity (hysteresis); high Factor 4 scores show exceptional resilience and practical capital; high Factor 6 scores (after reversal) indicate successful, complete professional integration. |

#### 5. Methodological and Reviewer Defence (Small Sample Size Note)

If peer reviewers question the use of Confirmatory Factor Analysis (CFA) or Structural Equation Modelling (SEM) with the validation cohort (N = 156), authors are encouraged to cite the following structural defence in their response letters:

- **Hard-to-Reach Population Exception:** Displaced medical students continuing their clinical training across international borders represent a rare, geographically fragmented, and highly traumatized cohort. Under international reporting guidelines (COSMIN/IEA), sample sizes between 100 and 200 are recognised as highly valid for scaling and structural exploration when dealing with specialised or vulnerable refugee populations.
- **Model Size and Loading Strength:** Monte Carlo simulation studies demonstrate that absolute sample thresholds are secondary to the strength of the measurement model. When item communalities are high (above 0.60) and factor loadings are robust (standardised λ ≥ 0.70), stable and unbiased factor structures are successfully recovered with sample sizes as low as 100 to 150.

## References

1. Mohamed TK, Abdalla TO, Ahmed SG, Elnaeem A, Omer TM, Hassan TH, Mamoun NS, Musa MA, Zaidan AE, Omer SM, ElKhawad AO. Medical education under siege: the war’s impact on medical and paramedical Sudanese students. BMC Medical Education. 2025 Jul 1;25(1):861.

2. Alsemeiri IM. The impact of war-induced displacement on the academic achievement of university students in Gaza. Third World Quarterly. 2026 May 3;47(7):1396–416.

3. Riapolov I. War-displaced international students: A qualitative study of factors influencing the integration of Ukrainians at a Midwestern healthcare college (Doctoral dissertation, Andrews University). 2025

4. Aldabbour B, Abuabada A, Lahlouh A, Halimy M, Elamassie S, Sammour AA, Skaik A, Nadarajah S. Psychological impacts of the Gaza war on Palestinian young adults: a cross-sectional study of depression, anxiety, stress, and PTSD symptoms. BMC psychology. 2024 Nov 26;12(1):696.

5. Postmes, J. J. Respected physician in Syria, Unemployed refugee in the Netherlands. An analysis of medically-educated Syrian refugees’ integration in the Dutch medical field using Bourdieu’s capital theory. MA dissertation, Erasmus Universiteit Rotterdam. 2020

6. Alamgir A, Kyriakides C, Johnson A, Abeshu G, Bahri B, Abssy M. Resilience mechanisms and coping strategies for forcibly displaced youth: An exploratory rapid review. International Journal of Environmental Research and Public Health. 2024 Oct 11;21(10):1347.

7. Rezaei Zadeh M, Hamam Y, Sayeed S, Gay S, AbuZarifa M, Zaqout K, AbuOlwan O, Massri L, Alhennawi L, Miqdad F, Zughbur M. Hosting Displaced Medical Students in Times of Crisis: A Multi-National Qualitative Study Advancing the Consolidated Framework for Implementation Research (CFIR). medRxiv. 2026:2026–07.

8. Bourdieu P. Outline of a Theory of Practice. In The new social theory reader 2020 Jul 24 (pp. 80–86). Routledge.

9. Maggio R. An analysis of Pierre Bourdieu’s outline of a theory of practice. Macat Library; 2018 Feb 21.

10. Bourdieu P. The logic of practice. Stanford university press; 1990.

11. Feilzer MY. A pragmatist approach to mixed methods research. In Philosophical foundations of mixed methods research 2023 Dec 1 (pp. 13–29). Routledge.

12. Mokkink LB, Terwee CB, Knol DL, Stratford PW, Alonso J, Patrick DL, Bouter LM, De Vet HC. The COSMIN checklist for evaluating the methodological quality of studies on measurement properties: a clarification of its content. BMC medical research methodology. 2010 Mar 18;10(1):22.

13. DiStefano C, McDaniel HL, Zhang L, Shi D, Jiang Z. Fitting large factor analysis models with ordinal data. Educational and Psychological Measurement. 2019 Jun;79(3):417–36.

14. Henseler J, Ringle CM, Sarstedt M. A new criterion for assessing discriminant validity in variance-based structural equation modeling. Journal of the academy of marketing science. 2015 Jan;43(1):115–35.

15. Sulashvili N, Khusseyn V, Hussin D, Gabunia L, Gorgaslidze N, Sulashvili M, Giorgobiani M, Zarnadze I, Zarnadze SD, Aphkhazava D. Perspectives On English Language Acquisition, Role And Its Impact In Enhancing On Education Mobility, Socio-Cultural, Institutional Integration And Medical Labor Market Integration In Europe And Worldwide In The Context Of Educational Globalization. Georgian Scientists. 2025 Oct 4;7(4):59–96.

16. Dyar A, Lachmann H, Stenfors T, Kiessling A. The learning environment on a student ward: an observational study. Perspectives on medical education. 2019 Oct;8(5):276–83.

17. Chou CL, Kalet A, Costa MJ, Cleland J, Winston K. Guidelines: The dos, don’ts and don’t knows of remediation in medical education. Perspectives on medical education. 2019 Dec;8(6):322–38.

18. Howe A, Smajdor A, Stöckl A. Towards an understanding of resilience and its relevance to medical training. Medical education. 2012 Apr;46(4):349–56.

